# Adiposity and metabolic health in Asian populations: An epidemiological study using Dual X-Ray Absorptiometry

**DOI:** 10.1101/2023.09.26.23296180

**Authors:** Theresia Mina, Xie Wubin, Dorrain Low Yan Wen, Wang Xiao Yan, Benjamin Lam Chih Chiang, Nilanjana Sadhu, Ng Hong Kiat, Nur-Azizah Aziz, Terry Yoke Yin Tong, Kerk Swat Kim, Choo Wee Lin, Low Guo Liang, Halimah Ibrahim, Lim Liming, Gervais Wansaicheong, Rinkoo Dalan, Yew Yik Weng, Paul Elliott, Elio Riboli, Marie Loh Chiew Shia, Joanne Ngeow Yuen Yie, Lee Eng Sing, Jimmy Lee Chee Keong, James Best, John Chambers

**Affiliations:** Nanyang Technological University Lee Kong Chian School of Medicine, Level 18 Clinical Sciences Building, 11 Mandalay Road, Singapore 308232; Khoo Teck Puat Hospital, Integrated Care for Obesity & Diabetes, 90 Yishun Central, Singapore 768828; Department of Diagnostic Radiology, Tan Tock Seng Hospital, 11 Jalan Tan Tock Seng, Singapore 308433; Department of Endocrinology, Tan Tock Seng Hospital, 11 Jalan Tan Tock Seng, Singapore 308433; National Skin Centre, Research Division, 1 Mandalay Rd, Singapore 308205; Department of Epidemiology and Biostatistics, School of Public Health, Imperial College London, 152 Medical School, St Mary’s Campus, London, United Kingdom W2 1NY; Division of Medical Oncology, National Cancer Centre, 11 Hospital Drive Singapore 169610; Clinical Research Unit, National Healthcare Group Polyclinic, 3 Fusionopolis Link, Nexus@one-north, #05-10, Singapore 138543; Research Division, Institute of Mental Health, Singapore 539747; Melbourne Medical School, University of Melbourne, Grattan Street, Parkville, Victoria, Australia 3010

## Abstract

**Background:** Diabetes, cardiovascular disease, and related cardiometabolic disturbances are increasing rapidly in the Asia-Pacific region. We investigated the contribution of excess adiposity, a key determinant of diabetes and cardiovascular risk, to unfavourable cardiometabolic profiles amongst Asian ethnic subgroups.

**Methods:** The Health for Life in Singapore (HELIOS) Study is a population-based cohort comprising multi-ethnic Asian men and women living in Singapore, aged 30-84 years. We analyzed data from 9,067 participants who had assessment of body composition by Dual X-Ray Absorptiometry (DEXA) and metabolic characterization. We tested the relationship of BMI and visceral Fat Mass Index (vFMI) on cardiometabolic phenotypes (glycemic indices, lipid levels, and blood pressure), disease outcomes (diabetes, hypercholesterolemia, and hypertension), and metabolic syndrome score with multivariate regression analyses.

**Findings:** Participants were 59.6% female, with mean (SD) age 52.8 (11.8) years. The prevalence of diabetes, hypercholesterolemia, and hypertension was 8.3%, 29% and 18.0%, respectively. Malay and Indian participants had 3-4 folds higher odds of obesity and diabetes, and showed adverse metabolic and adiposity profiles, compared to Chinese participants. Excess adiposity contributed to all adverse cardiometabolic health indices including diabetes (P<0.001). However, while vFMI explained the differences in triglycerides and blood pressure between the Asian ethnic groups, increased vFMI did not explain higher glucose levels, reduced insulin sensitivity and risk of diabetes amongst Indian participants.

**Interpretation:** Visceral adiposity is an independent risk factor for metabolic disease in Asian populations, and accounts for a large fraction of diabetes cases in each of the ethnic groups studied. However, the variation in insulin resistance and diabetes risk between Asian subgroups is not consistently explained by adiposity, indicating an important role for additional mechanisms underlying the susceptibility to cardiometabolic disease in Asian populations.

**Funding:** Nanyang Technological University—the Lee Kong Chian School of Medicine, National Healthcare Group, National Medical Research Council, Singapore.

**Research in context:** 

**Evidence before this study:** We searched Embase and MEDLINE using MeSH terms and respective alternative terms for [“body fat distribution” OR “visceral adiposity” OR “diagnostic imaging”] and [“metabolic syndrome” OR “diabetes mellitus” OR “hypertension” OR “hyperlipidemia” OR *all corresponding phenotypes*] from 1946 till 7^th^ August 2023 and identified 456 relevant studies. Overall, there have been substantial attempts to characterize the impact of adiposity quantified with imaging techniques on cardiometabolic health. However, most works focused on validating novel adiposity indices (such as body shape index) or metabolic biomarkers (such as cytokines), and rarely provided insights on the contribution of excess visceral adiposity across cardiometabolic phenotypes. Some investigations focused on delineating the effect of various fat depots in the viscera on insulin resistance. Very few studies evaluated health disparity across populations; Nazare et al. characterized the impact of visceral vs. subcutaneous fat measured using Computed Tomography on various cardiometabolic outcomes across major ethnic groups in United States. In summary, it remains unclear how visceral adiposity contributes to differences in cardiometabolic health burden across large Asian ethnic groups.

**Added value of this study:** Our multi-ethnic population cohort (n=9,067) included standardized assessments of people of Chinese, Malay, and Indian ancestries living in shared environment, bringing relevance to a wide spectrum of global Asian diaspora. We used the whole-body DEXA-based quantification of visceral fat mass which enables separate assessments of visceral adiposity and overall body fat. We show that there are major differences in adiposity and metabolic health between the Chinese, Malay, and Indian Asian people we studied, and that adiposity makes an important contribution to metabolic health in all three of these Asian ethnic subgroups. However, we also show that excess visceral adiposity only partially explains the difference in diabetes, insulin resistance and related metabolic disturbances between major Asian ethnic subgroups, indicating the presence of additional pathophysiological processes that remain to be identified.

**Implications of all the available evidence:** Excess visceral adiposity is an important contributor to cardiovascular and metabolic health in Asian populations. Strategies to reduce excess adiposity, in particular visceral fat, in Malay and Indian subgroups offer opportunities for major improvements in cardiometabolic health in Asian people, who account for ∼60% of the global population. The difference in diabetes, insulin resistance and related metabolic disturbances between major Asian ethnic subgroups remains unexplained, providing the motivation for further research to identify additional pathophysiological processes underlying these leading global diseases.

## 1. Introduction

Diabetes and cardiovascular disease (CVD) are closely interlinked chronic diseases that are leading causes of morbidity and mortality worldwide. Over the last 3 decades, the global distribution of diabetes and CVD has been progressively shifting away from Europe and North America towards the emerging market economies of the Asia-Pacific region^1–3^. There are already 296 million people living with diabetes in the Asia-Pacific region^4^, and this number is projected to increase by a further 40% to 412 million by 2040^5^. The number of deaths caused annually by CVD in Asia has also increased from 5 million in 1990 to 9 million in 2019^6^. There is an urgent need to better understand the determinants of cardiometabolic risk in Asian individuals, as part of national and global efforts aimed at prevention of diabetes and CVD.

While cardiovascular and metabolic disease are increasing across the Asia-Pacific region, the burden of disease differs between the respective countries and ethnic subgroups of Asia^1–3^. Cross-sectional and prospective population studies have consistently shown that the risks for diabetes and CVD are highest amongst Asian populations living in urban environments^4,5^, and that disease risks are also raised amongst people of South Asian background compared to their East Asian counterparts^7^. In contrast, the burden of hypertension and stroke are higher in East Asians^3^. The mechanisms underlying this heterogeneity in disease outcomes remain unknown; this represents a major obstacle to effective risk stratification and preventative interventions for Asian communities.

Diabetes and CVD are complex multi-factorial diseases, with important contributions from behavioral, environmental, and heritable factors. Genetic association studies have identified hundreds of common and rare genetic variants that contribute to disease and disease-related endophenotypes^8–10^. However, these adverse genetic exposures are typically shared between populations, and do not explain the high risk of disease in Asian individuals^7–9^. In contrast, observational studies have consistently documented an increasing burden of adiposity amongst Asian populations. BMI and waist circumference (a marker of visceral adiposity) have both increased in Asian populations over the last 2 decades,^11^ although the magnitude varies between Asian subgroups. In particular, visceral adiposity is closely associated with the presence of insulin resistance, atherogenic dyslipidemia, raised blood pressure and immune activation, and is causally linked to the development of both diabetes^12^ and CVD^13^. These observations raise the possibility that increasing adiposity is a key determinant of the adverse cardiovascular and metabolic profiles amongst Asian populations, and of the heterogeneity in cardiometabolic health between Asians in different settings^14^.

To address this critical question, we carried out comprehensive evaluation of a large multi-ethnic Asian population cohort, including assessment of body fat composition by DEXA, to i) quantify the relationships between adiposity and risks for metabolic and cardiovascular phenotypes, amongst people of Chinese, Malay, and Indian background, and ii) test the hypothesis that visceral adiposity explains the differences in cardiometabolic health observed between these three major Asian ethnic subgroups.

## 2. Materials & Methods

### 2.1. General study information

The Health for Life in Singapore study (HELIOS, IRB approval by Nanyang Technological University: IRB-2016-11-030, www.healthforlife.sg) is an observational population cohort study comprising the multi-ethnic Asian population of Singapore, a country with three primary ethnic groups: Chinese [76%], Malay [15%] and Indian [9%] Asians. We included men and women aged 30-84 years old and excluded pregnant or breastfeeding women, individuals with major illness/ surgery, or participation in pharmaceutical trials within the past month, and cancer treatment in the past year. All participants were recruited from the general population to ensure diversity in ethnicity and socio-economic background.

### 2.2. Characterization of participants

Participants completed a comprehensive, structured assessment including questionnaires, physiological measures, and imaging assessments. Biological samples were collected in the fasted state (≥8 hours). Participants’ self-reported ethnicity was recorded based on their Singapore National Registration Identity Card, along with country of birth, nationality, religion, and primary language. Based on these data, 9,879 participants (98.7%) were classified into one of three ethnic groups: i. Chinese and other East Asian (‘Chinese’); ii. Malay and other South-East Asian (‘Malay’) or iii. Indian and other South Asian (‘Indian’). We have previously shown that this classification of ethnicity aligns closely to ancestry assessed objectively by genetic information^15^. The remaining 125 individuals (1.3%) were from non-Asian or mixed population groups and were excluded from the present analysis. Socio-economic status (highest level of education and household income), lifestyle information (cigarette smoking and alcohol intake), and health history were assessed as part of comprehensive health and lifestyle questionnaires. The highest level of education was converted to total years of education^16^. Information on participants’ medication was obtained through structured nurse interview. All generic and proprietary names of prescribed medication for diabetes, hypercholesterolemia, and hypertension were recorded.

Resting blood pressure was measured 3 times using automated blood pressure monitoring devices. HDL cholesterol (HDL-C), LDL cholesterol (LDL-C), triglycerides, and glucose were measured from fasting blood samples by the accredited laboratory (QuestLab, Singapore, SAC–SINGLAS ISO 15189:2012). Fasting insulin was measured with immunoassays using the ADVIA Centaur XP Immunoassay System (Siemens Healthcare, Erlangen, Germany). Homeostatic Model Assessment for Insulin Resistance (HOMA-IR) = insulin [mIU/L]*glucose [mmol/L])/22.15, and HOMA for β-cell function (HOMA-%B) = (20*insulin [mIU/L])/(glucose[mmol/L]-3.5) ^17^. Diabetes was defined as: i. self-reported diagnosis of diabetes and on pharmacological treatment for diabetes, or ii. fasting glucose ≥7.0mmol/L^4,5,18^. Hypertension was defined as: i. self-reported diagnosis of hypertension and on blood pressure lowering medication, or ii. blood pressure ≥140/90 mmHg^3,18^. Hypercholesterolemia was defined as: i. self-reported diagnosis of high cholesterol and on lipid lowering treatment, or ii. LDL-cholesterol ≥4.1 mmol/L^18^.

Body height and weight were measured to calculate BMI. Waist circumference was measured 3 times and averaged. Visceral Adipose Tissue (VAT, *Kg*) was assessed by DEXA whole-body scans, performed using Horizon^TM^ W densitometer (S/N 30052M, Hologic, Massachusetts, USA, QDR software version 13.6.0.5). VAT was estimated by subtracting subcutaneous fat from total fat in the L4 region using an algorithm validated against CT scan ^19^ and MRI ^20^. VAT was converted into common units (Kg) and normalized by height^2^ to derive visceral Fat Mass Index (vFMI, *Kg/m*^2^), as previously reported^16^. Overweight and Obesity are defined as BMI ≥23.0 Kg/m^2^ and >27.5 Kg/m^2^, respectively. Central obesity is defined as ≥90 cm and ≥80 cm for male and female, respectively^21^.

### 2.3. Statistical analysis

R version 4.2.2 was used for statistical analysis and illustrations (**S Methods 1**). Significance was evaluated at the 0.05 level. In this study, we defined 3 datasets out of 10,004 HELIOS participants screened between April 2018 to January 2022: i) all eligible participants (n=9,067); ii) participants not on medications for diabetes, hypercholesterolemia, or hypertension (n=6,807); and iii) repeat participants recalled within 20 months (n=230) (**S Methods 2,** CONSORT diagram, **S Figure 1**). Outliers were identified amongst continuous variables and were verified against source documents. No observations were excluded after correction. Triglycerides, HDL-C, glucose, insulin, HOMA-IR, and HOMA-%B were *Ln*-transformed to achieve normal distributions.

Initial descriptive analyses of demographic, metabolic, and adiposity variables were performed across ethnic groups using Chi-square test and one-way ANOVA (for categorical and continuous variables, respectively).

We next compared the metabolic and adiposity phenotypes across ethnicity using General Linear Modelling. We calculated sex-adjusted Estimated Marginal Means (EMM) of glucose, insulin, HOMA-IR, HOMA-%B, triglycerides, HDL-C, systolic and diastolic blood pressure (SBP and DBP, respectively), BMI, waist circumference, and vFMI across ethnicity, and age-by-decades. To assess the contribution of adiposity to metabolic health, we performed i) multivariate linear regressions using BMI or vFMI as predictors for glucose, insulin, HOMA-IR, HOMA-%B, triglycerides, HDL-C, SBP, and DBP, and ii) logistic regressions with diabetes, hypercholesterolemia, and hypertension as outcomes. All regression models were adjusted for sex, age, and ethnicity. Population attributable disease risks to ethnic differences in obesity and central obesity were calculated across ethnicity, adjusted for sex and age.

Subsequently, to evaluate the contribution of excess adiposity to ethnic differences in metabolic health, we calculated the differences in EMM between ethnicities (ΔEMM) of the metabolic phenotypes across 3 multivariable regression models: a) adjusted for sex and age only, b) model a + BMI, and c) model a + vFMI. Chinese ethnicity was the reference group. As sensitivity analyses, we repeated the ethnic-specific regression analysis in i) all eligible datasets inclusive of participants with medication, or ii) using the untransformed raw data, or iii) after accounting for regression dilution bias, or iv) after further adjusting for lifestyle and socioeconomic status (cigarette smoking, alcohol intake, total year of education and household income). Regression dilution bias was estimated by calculating correction factor 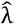, where corrected beta 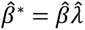 Correction factor was derived from two-way mixed effects IntraClass Coefficient (ICC) for absolute agreement between individuals who attended the repeat visit (n=230), where 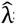 = 1/ICC^22^. **S Methods 3** provides further details on the mathematical equations required to derive 95% Confidence Interval (CI) of 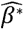.

To further examine the relationships between adiposity and metabolic disturbances in Asian populations, we evaluated ethnic difference in the contribution of excess adiposity towards metabolic syndrome score. To formulate the score, we selected non-collinear, representative metabolic measures using a hierarchical clustering of the inter-correlation matrix of the measures. Since adiposity is one of the metabolic syndrome components^21^, we excluded adiposity from our metabolic syndrome score derivation. We then performed Principal Components (PCs) analysis of these representative measures and evaluated factor loadings of each component across PCs, and the corresponding total sample variance (adjusted R-squared). We selected the first PC (PC1) as the primary metabolic syndrome score. We then evaluated the contribution of excess adiposity to the ethnic-specific ΔEMM of PC1 across 3 multivariable regression models as previously described.

## 3. Results

### Characteristics of the study participants

There were 10,004 people recruited to the HELIOS cohort, amongst whom 9,067 had complete data required for the current analysis (CONSORT Table, **S Figure 1**). These 9,067 participants were 59.6% female, with mean ± SD age 52.8±11.8 years (**S Table 1**). Self-reported ethnicity was 69% Chinese, 13% Malay, and 18% Indian (**S Table 1**). The prevalence of obesity, diabetes, hypercholesterolemia, and hypertension was 12.2%, 8.3%, 29%, and 18.0%, respectively in the study population. These distributions approximate those reported in national surveys of the general population (**S Table 1**).

There were major differences in the burden of obesity and cardiometabolic disease between the Asian ethnic sub-groups (**Table 1**). In particular, obesity and diabetes were 3 to 4-fold more common in Malay and Indian participants, compared to Chinese participants (obesity: 44.6% and 38.8%, vs 11.5% respectively; diabetes: 13.6% and 17.7%, vs 4.9% respectively; both P<2.2 x10^-16^, **Figure 1**). BMI, waist circumference and vFMI were all higher, while central obesity, hypercholesterolemia and hypertension were more common, amongst Malay and Indian participants, compared to Chinese participants (P<0.001, **Figure 1**).

**Table 1.** The demographic, metabolic, and adiposity characteristics of participants with no medication for diabetes, hypertension, and hypercholesterolemia. , n=6,807. Geometric mean ± SD are presented for glucose, insulin, HOMA-IR, HOMA-%B, triglycerides, and HDL-C.

| General and socio-demographic factors, categorical n (%) unless indicated |  | Chinese (n=4,789) | Malay (n=879) | Indian (n=1,139) | P |
| --- | --- | --- | --- | --- | --- |
| Age (years) mean ± SD |  | 50.59 ± 11.63 | 48.67 ± 10.49 | 49.7 ± 10.47 | 6.2 × 10<sup>-6</sup> |
| Sex | Male | 1,760 (36.8) | 300 (34.1) | 536 (47.1) | 3.2 × 10<sup>-11</sup> |
|  | Female | 3,029 (63.2) | 579 (65.9) | 603 (52.9) |  |
| Cigarette smoking | Never smoke | 4,147 (86.6) | 585 (66.6) | 895 (78.6) | <2 × 10<sup>-16</sup> |
|  | Ex-smoker | 343 (7.2) | 117 (13.3) | 103 (9.0) |  |
|  | Active smoker | 299 (6.2) | 177 (20.1) | 141 (12.4) |  |
| Alcohol intake | Never or special occasion | 3,685 (77.1) | 852 (97.0) | 851 (74.7) | <2 × 10<sup>-16</sup> |
|  | 1-2/month or 1-2/week | 864 (18.1) | 20 (2.3) | 227 (19.9) |  |
|  | 3-4/week or daily | 229 (4.8) | 6 (0.7) | 61 (5.4) |  |
| Total year of education, mean (SD) |  | 14.42 ± 3.18 | 12.35 ± 3.17 | 14.1 ± 3.44 | <2 × 10<sup>-16</sup> |
| Household income (SGD\$) | <3,999 | 964 (20.1) | 316 (35.9) | 325 (28.5) | <2 × 10<sup>-16</sup> |
|  | 4,000-9,999 | 1,789 (37.4) | 361 (41.1) | 465 (40.8) |  |
|  | >=10,000 | 1,389 (29.0) | 91 (10.4) | 225 (19.8) |  |
|  | Do not know | 72 (1.5) | 19 (2.2) | 17 (1.5) |  |
|  | Prefer not to answer | 140 (12.0) | 52 (10.5) | 48 (9.4) |  |
| Adiposity and metabolic components, mean ± SD, unless indicated |  |  |  |  |  |
| Glucose (mmol/L) |  | 4.78 ± 0.56 | 4.9 ± 1.05 | 5.03 ± 1.11 | <2 × 10<sup>-16</sup> |
| Insulin (μIU/mL) |  | 7.15 ± 5.57 | 9.38 ± 7.38 | 11.37 ± 9.07 | <2 × 10<sup>-16</sup> |
| HOMA-IR |  | 1.52 ± 1.32 | 2.04 ± 1.93 | 2.54 ± 2.44 | <2 × 10<sup>-16</sup> |
| HOMA-%B |  | 117.38 ± 87.85 | 150.47 ± 136.26 | 164.38 ± 147.13 | <2 × 10<sup>-16</sup> |
| Triglycerides (mmol/L) |  | 1.01 ± 0.58 | 1.17 ± 0.71 | 1.19 ± 0.67 | <2 × 10<sup>-16</sup> |
| HDL-C (mmol/L) |  | 1.59 ± 0.46 | 1.4 ± 0.39 | 1.28 ± 0.37 | <2 × 10<sup>-16</sup> |
| Systolic blood pressure (mmHg) |  | 118.05 ± 17.35 | 119.64 ± 17.26 | 119.21 ± 17.29 | 0.0119 |
| Diastolic blood pressure (mmHg) |  | 69.69 ± 10.53 | 72.5 ± 10.34 | 73.16 ± 10.89 | <2 × 10<sup>-16</sup> |
| BMI (Kg/m<sup>2</sup>) |  | 23.22 ± 3.66 | 27.57 ± 4.92 | 26.72 ± 4.70 | <2 × 10<sup>-16</sup> |
| BMI ≥27.5 Kg/m2, n (%) |  | 553 (11.5) | 392 (44.6) | 442 (38.8) | <2 × 10<sup>-16</sup> |
| Waist circumference (cm) |  | 78.99 ± 10.06 | 86.03 ± 11.42 | 88.33 ± 11.64 | <2 × 10<sup>-16</sup> |
| Waist circumference ≥90 cm and ≥80 cm for male and female, respectively, n (%) |  | 1,312 (27.4) | 517 (58.8) | 698 (61.3) | <2 × 10<sup>-16</sup> |
| Visceral FMI (Kg/m<sup>2</sup>) |  | 0.2 ± 0.09 | 0.26 ± 0.11 | 0.27 ± 0.1 | <2 × 10<sup>-16</sup> |

**Figure 1.**
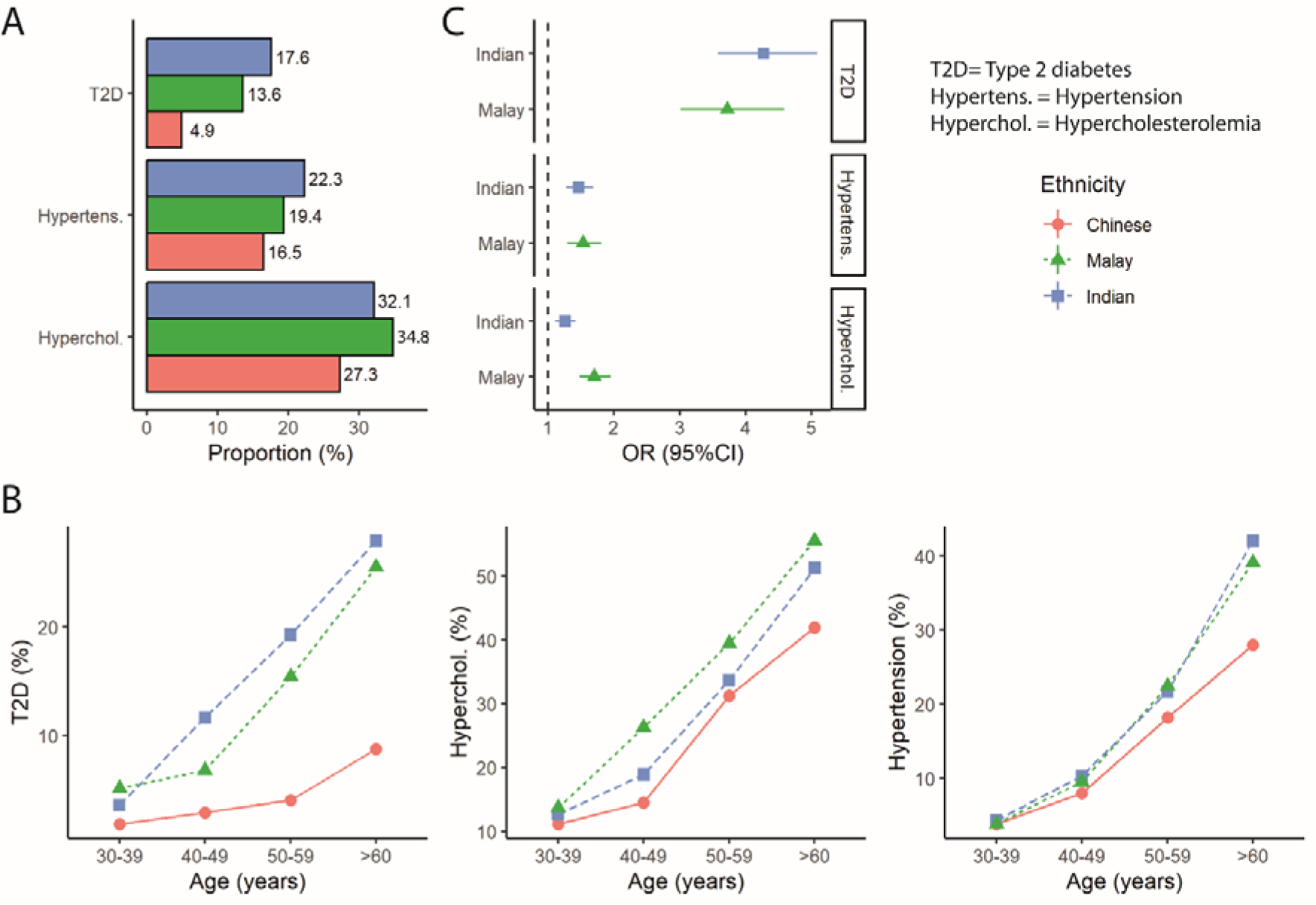
Metabolic disease status across age according to ethnicity, n=9,067. **A)** Overall disease proportion; **B)** Disease proportion across age by decades; **C)** The odds of disease by ethnicity, with Chinese as reference, adjusted for sex and age. Diabetes was defined as glucose ≥ 7.0 mmol/L or on treatment. Hypertension was defined as BP ≥140/90 mmHg or on treatment. Hypercholesterolemia was defined as LDL-C ≥4.1 mmol/L or on treatment.

### Adiposity is a major risk factor for cardiovascular and metabolic disease in Asian populations

Amongst the 9,067 participants included in the study, raised BMI, waist circumference and vFMI were each associated with an increased risk for diabetes, hypertension, and hypercholesterolemia (**S Figure 2A**, 95% CI of all OR>1, all P≤6.7×10^-37^). The relationship of adiposity with adverse metabolic outcomes appeared stronger for vFMI compared to BMI (**S Figure 2A**). Numerically, a 1SD increase in vFMI was associated with a 1.9-fold increased risk of diabetes, a 1.8-fold risk for hypertension, and a 1.4-fold risk for hypercholesterolemia. Based on the observed relationship between adiposity and diabetes, the adjusted population risks for diabetes attributable to obesity, overweight, and central obesity were 16.0, 30.8% and 40.8% respectively (**S Table 2**, **Figure 2C**). Findings were similar in analyses within each of the Asian ethnic groups (, **Figure 2C, S Table 2**). Population risks for hypertension and hypercholesterolemia attributable to central obesity were also high (**S Table 2,, Figure 2C**).

**Figure 2.**
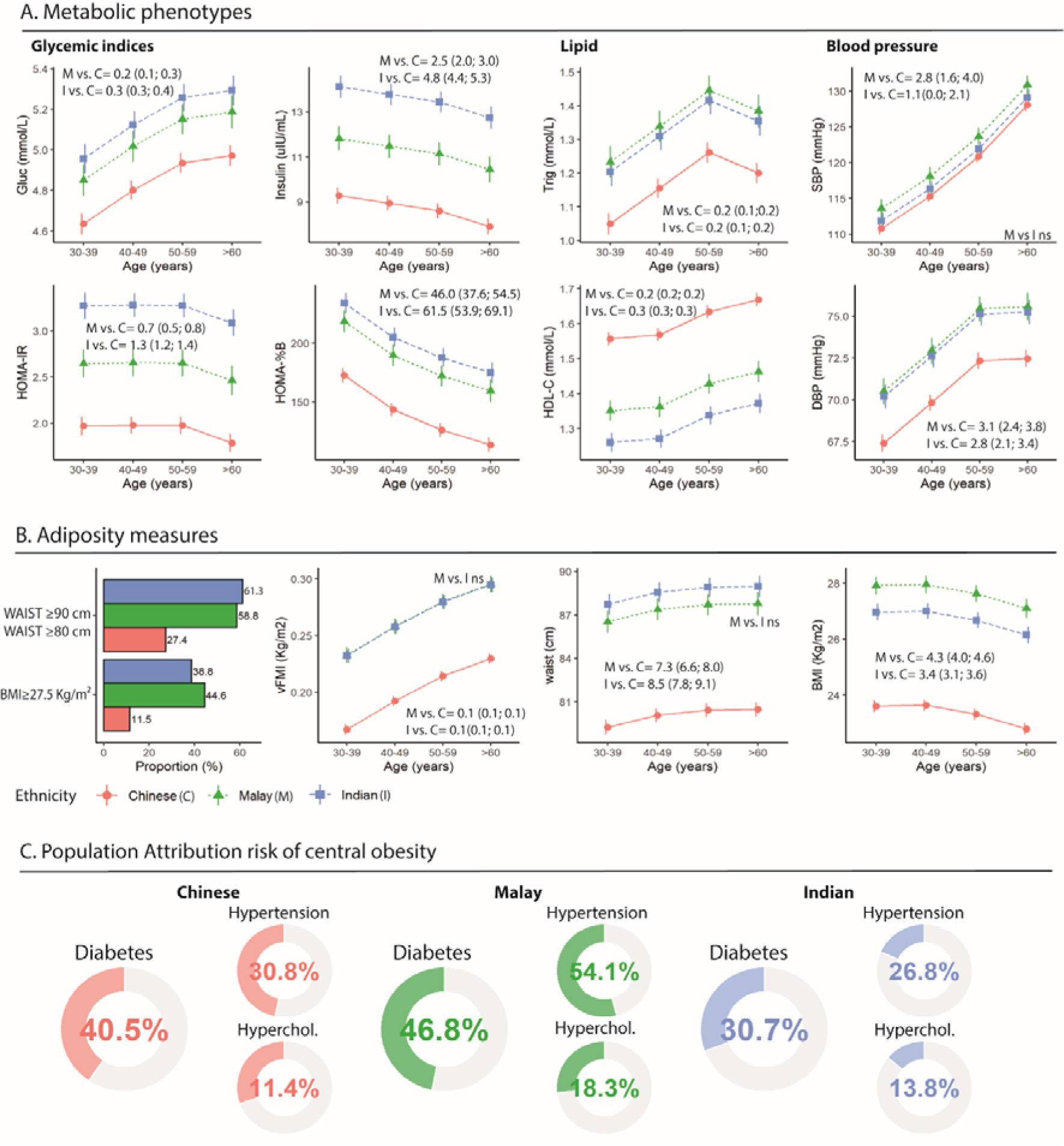
Metabolic phenotypes and adiposity across age and ethnicity in people with no medications for diabetes, hypertension, and hypercholesterolemia, n= 6,807. X-axis is age in years by decades. **A)** Metabolic phenotypes including (L-R) glycemic indices, lipid measures, and blood pressure; **B)** Adiposity indices. The estimated marginal means ±SEM in sub-figure A) and B) were derived from general linear modelling adjusted for sex. Ethnic comparisons were all p<0.001 except as indicated in the figure (ns=not significant). Each diagram includes 2 sets of numbers, which are ΔEMM between M vs. C (Malay vs. Chinese) or I vs. C (Indian vs. Chinese). **C)** Adjusted population attribution risk percentage for central obesity in all available dataset (**n=9067)**, defined as≥90 cm and ≥80 cm for male and female, respectively. **Abbreviations (A-Z):** BMI= body mass index; DBP= diastolic blood pressure; Gluc= Glucose; HDL-C= High-density lipoprotein cholesterol; HOMA-%B= Homeostatic model assessment (HOMA) of β-cell function; HOMA-IR =Homeostatic Model Assessment for Insulin Resistance; hyperchol. = hypercholesterolemia; **S**BP= systolic blood pressure; Trig= Triglycerides; vFMI= visceral Fat Mass Index; waist= waist circumference (men ≥90cm, women ≥80cm).

Amongst participants with normal BMI (<23 kg/m^2^), the prevalence of T2D was 3.0% in Chinese, 3.7% in Malay, but 18.7% in Indian Asians (P<2.2×10^-16^, **S Table 3**). The 3.7-fold increased risk of T2D in Malay individuals was greatly reduced after adjusting for high levels of adiposity compared to Chinese participants (OR for T2D: 2.0, 2.3 and 2.2 after adjustment for BMI, waist circumference or vFMI respectively, **S Table 4**). In contrast, the high prevalence of T2D amongst Indian Asians compared to Chinese was only partially explained by raised BMI, waist circumference or vFMI (OR for T2D: 3.2, 2.8 and 3.0 after adjustment for respective adiposity traits).

### Heterogeneity in insulin resistance and related disturbances between Asian populations

There were 6,807 participants who were not on treatment for diabetes, hypercholesterolemia, or hypertension. Their cardiometabolic profiles are summarized in **Figure 2A and Table 1**. Malay and Indian participants had higher BMI and waist circumference, compared to Chinese participants (**Table 1**, **Figure 2B**, BMI: 27.6±4.9 and 26.7±4.7 vs. 23.2±3.7 Kg/m^2^; waist: 86.0±11.4 and 88.3±11.6 vs. 79.0±10.1 cm, respectively, both P<2.2 x10^-16^). DEXA confirmed increased visceral adiposity in Malay and Indian participants, compared to Chinese (vFMI: 0.26±0.11 and 0.27±0.1 vs. 0.20±0.09 Kg/m^2^ respectively, P<2.2x10^-16^). Both Malay and Indian participants also had markedly higher fasting glucose, insulin, HOMA-IR, triglycerides, and lower HDL-C (all P<2.2 ×10^-16^; **Figure 2**). The adverse metabolic profiles of Malay and Indian individuals were evident across all ages studied **(Figure 2B**). SBP and DBP were higher in Malay and Indian participants compared to Chinese participants (P<0.05), although the difference was smaller than observed for metabolic traits.

### Does adiposity explain the difference in insulin resistance and related metabolic disturbances between Asian ethnic subgroups?

Amongst the 6,807 participants not on drug treatment for cardiometabolic disease, raised BMI, waist circumference and visceral adiposity were strongly associated with increased fasting glucose, insulin, HOMA-IR, blood pressure, triglycerides, and with lower HDL-C, overall (P<2.2×10^-16^, **S Figure 2B**), and in each of the ethnic groups (P<2.2×10^-16^, **S Figure 2C**). The relationship of adiposity with adverse metabolic outcomes was again stronger for visceral fat compared to BMI (**S Figure 2B**). Numerically, a 1SD increase in vFMI was associated with an 0.28 SD increase in ln-glucose, and an 0.63 SD increase in ln-HOMA-IR (all P<2.2×10^-16^).

We used multivariable analyses to test the contribution of visceral adiposity to the differences in cardiometabolic traits between Chinese, Malay, and Indian participants. *Glucose and insulin action.* Visceral adiposity did not explain the high levels of glucose, insulin, and HOMA-IR amongst Indian, compared to Chinese participants (**Figure 3, S Table 5**). In particular, HOMA-IR remained elevated in Indian participants despite adjusting for visceral adiposity (Difference [95% confidence interval] in z-ln-HOMA-IR between Indian vs Chinese participants: i. Unadjusted: 0.74 [0.69 to 0.81]; ii. Adjusted for vFMI: 0.31 [0.26 to 0.36]; both P<2.9×10^-12^), or after adjustment for any other available measure of adiposity (**S Table 6**). In contrast, increased visceral adiposity fully explained these metabolic defects amongst Malay compared to Chinese participants. *Triglycerides and HDL-C*. We found that differences in visceral adiposity fully explained the variation in triglyceride concentrations between the Chinese, Malay, and Indian participants (**Figure 3, S Table 5)**. In contrast, visceral adiposity only partially explained the differences in HDL-C concentrations between the Asian subgroups (**Figure 3, S Table 5**). The difference in z-ln-HDL-C between Malay and Chinese participants was: i. unadjusted: 0.49 [0.43 to 0.56]; ii. adjusted for vFMI: 0.20 [0.14 to 0.26], both P<7.8×10^-11^), and between Indian vs Chinese was i. unadjusted: 0.75 [0.70 to 0.81]; ii. adjusted for vFMI: 0.47 [0.42 to 0.52], both P<2.9×10^-12^). *Blood pressure.* Visceral adiposity largely explained the differences in both SBP and DBP (**Figure 3, S Table 5**) between the ethnic groups.

**Figure 3.**
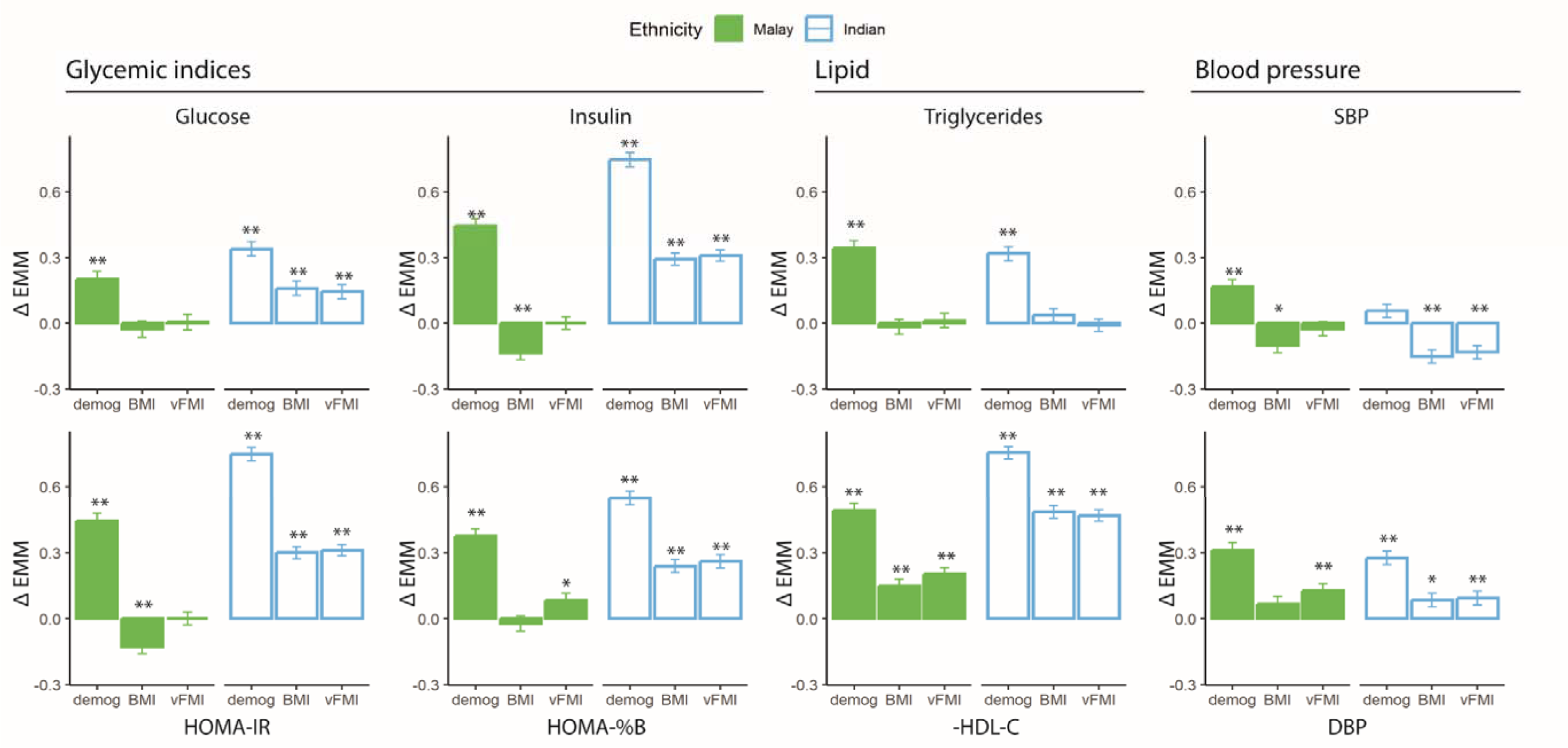
Contribution of excess visceral adiposity in metabolic health across ethnicity. All analyses were adjusted for sex and age, and were performed in participants without medication for diabetes, hypertension, and hyperlipidaemia, n**=** 6,807. Glucose, insulin, HOMA-IR, HOMA-%B, triglycerides and HDL-C were all ln-transformed and z-scored. Each bar represents Δ Estimated Marginal Means ±SEM of z-scores before and after adjusting for adiposity indices. ΔEMM for z-HDL-C was inversed to ease comparison. The 3 regression models were i) demog= adjusted for sex and age; ii) BMI=model i) + BMI; and iii) vFMI=model i) + vFMI. ** p<0.001 and * p <0.05 compared to Chinese participants as reference. **Abbreviations (A-Z):** BMI= body mass index; DBP= diastolic blood pressure; Gluc= Glucose; HDL-C= High-density lipoprotein cholesterol; HOMA-%B= Homeostatic model assessment (HOMA) of β-cell function; HOMA-IR=Homeostatic Model Assessment for Insulin Resistance; SBP= systolic blood pressure; vFMI= visceral Fat Mass Index. Fat mass estimated as BMI or vFMI does not explain differences between Indian and Chinese participants in glycemic indices and HDL-C levels.

Results for these primary analyses were similar when BMI was used as an alternate measure of adiposity, and in sensitivity analyses that included participants on drug treatment (**S Figure 3**), or when untransformed data were used (**S Figure 4**). These observations were not materially changed by adjustment for regression dilution (**S Table 7, S Figure 5**), or for socio-economic status (**S Figure 6**). To achieve the levels of insulin sensitivity equivalent to those observed in Chinese participants, we estimate that Malay and Indian participants would need to achieve a mean BMI 23.5 and 19.9 Kg/m^2^, respectively (**S Table 8**).

### Contribution of visceral adiposity to composite measures of metabolic performance

Finally, we used Principal Components Analysis (PCA) to generate composite scores that describe cardiometabolic phenotype amongst the Asian individuals in our study population. Hierarchical clustering of the correlation matrix amongst metabolic and adiposity parameters identified HDL-C, HOMA-IR, triglycerides, glucose, and DBP as representative non-adiposity metabolic parameters for inclusion in the PCA model (**S Figure 7**). PC1 alone explained 47% of total variance, and showed similar loadings across the underlying metabolic components (**Figure 4A**). The subsequent PCs were centered on the separate metabolic parameters, and cumulatively explained the remaining variance (**Figure 4A**).

**Figure 4.**
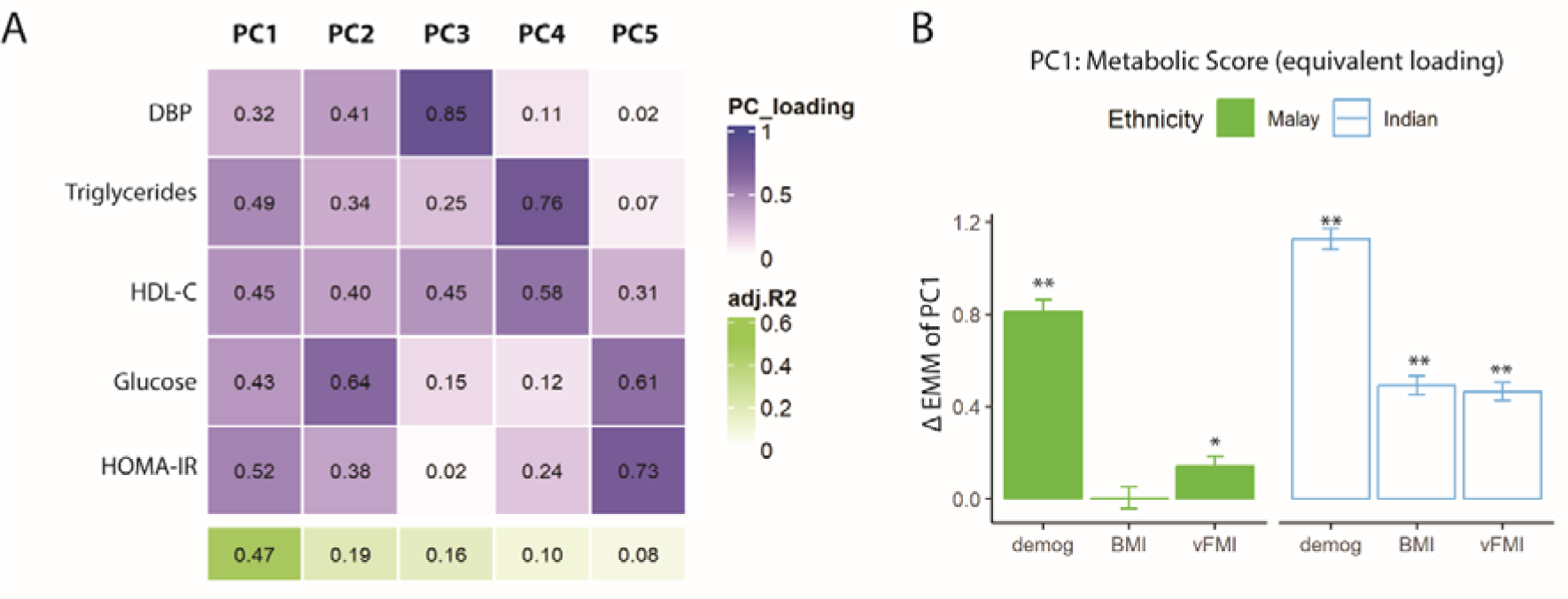
Contribution of visceral adiposity to metabolic variation in Asian populations. All analyses were adjusted for sex and age, and were performed in participants without medication for diabetes, hypertension, and hyperlipidaemia, n**=** 6,807. Glucose, HOMA-IR, triglycerides, and HDL-C were all ln-transformed and z-scored. **A)** Principal Component Analysis of representative non-adiposity metabolic phenotypes after dimensional reduction. Each column is one Principal Component (PC), with the corresponding adjusted R^2^. Each row represents factor loading across PCs. The first PC product was used to represent metabolic syndrome continuous score. **B)** Ethnic difference in metabolic syndrome (PC1) before and after adjusting for adiposity measures. Each bar represents Δ Estimated Marginal Means ±SEM of PC1 compared to Chinese participants as reference, x-axis indicates covariates (sex and age), with 1 additional adiposity parameter. ** p<0.001 and * p <0.05 compared to Chinese as reference**. Abbreviations (A-Z):** BMI= body mass index; DBP= diastolic blood pressure; Gluc= Glucose; HDL-C= High-density lipoprotein cholesterol; HOMA-IR=Homeostatic Model Assessment for Insulin Resistance; Trig= Triglycerides; vFMI= visceral Fat Mass Index. Adjustment for adiposity does not explain difference in metabolic syndrome between Indian and Chinese participants.

PC1 was higher in Malay and Indian participants, compared to Chinese participants (PC1 z-scores: Chinese, -0.18±0.02; Malay, 0.63±0.04; Indian, 0.95±0.04; P=2.9×10^-12^; **Figure 4B**). Visceral adiposity largely explained the difference in PC1 between Malay and Chinese participants (difference in PC1 z-score: i. unadjusted: 0.81 [0.71 to 0.91], P=2.9×10^-12^; ii. adjusted for vFMI: 0.14 [0.06 to 0.22], P=0.002; **Figure 4B**). In contrast, PC1 remained high amongst Indian participants after adjustment for visceral adiposity (difference in PC1 z score: i. unadjusted: 1.12 [1.03 to 1.22], P=2.9×10^-12^; ii. adjusted for vFMI: 0.46 [0.39 to 0.54], P=1.09×10^-12^). Visceral adiposity also did not consistently explain the differences in PC2-PC5 between the Asian ethnic subgroups (**S Figure 6**).

## Discussion

We demonstrate that adiposity, cardiovascular and metabolic profiles differ between people in Singapore of Chinese, Malay, and Indian Asian background, being most favorable amongst Chinese individuals. We confirm that adiposity is strongly associated with diabetes, insulin resistance and related metabolic disturbances overall, and in each of the Asian subgroups. We also showed that, however, adiposity only partially explains the wide differences in cardiometabolic health observed amongst the three Asian ethnic subgroups evaluated. In particular, while visceral adiposity accounts for most of the cardiometabolic disturbances in Malay compared to Chinese individuals, the high levels of insulin resistance and diabetes amongst Indian individuals were not fully explained by their increased adiposity. Our results have important implications for our understanding of the etiology of metabolic disease, and for interventions to improve metabolic health in Asian populations, who account for ∼60% of the global population.

Although cardiovascular and metabolic disease are increasing across the Asia-Pacific region, the burden of disease shows marked heterogeneity between geographic, socio-economic, and demographic settings. Identification of the mechanisms underlying these divergent cardiometabolic outcomes is a critical step towards implementation of effective risk stratification and preventative interventions for Asian communities. To address this need, we carried out a population-based study of men and women living in Singapore, from the Chinese, Malay, and Indian Asian communities. Participants underwent comprehensive characterization of cardiovascular and metabolic health, including quantification of body fat composition by DEXA. As our primary research question, we used the data to test the hypothesis that adiposity explains the differences in cardiometabolic health observed between the three major Asian ethnic subgroups evaluated.

We show that diabetes, insulin resistance, and related metabolic disturbances are more common in Malay and Indian participants, compared to people from the Chinese community, despite living in a shared geographic setting. In particular, the prevalence for diabetes was almost 4-fold higher amongst Malay and Indian participants compared to Chinese participants. There were also striking differences in adiposity between Asian ethnic subgroups. Mean levels for BMI, waist circumference and visceral adiposity were highest in Malay and Indian participants, with more than half of people from these ethnic backgrounds having obesity and/or central obesity. Our results are consistent with findings in Europe and North America which show that Indian and other South Asian populations have high levels of diabetes and central obesity, and reduced insulin sensitivity compared to Europeans and East Asians^7,23–27^, and provide further evidence of the disparate metabolic outcomes amongst global Asian populations.

Our analyses confirm the adverse impact of excess adiposity on cardiovascular and metabolic health. Increased adiposity was strongly associated with an increased risk for diabetes and hypertension, elevated insulin resistance and triglycerides, and with reduced HDL-C, independent of age and sex. The relationships were present in each of the Asian ethnic groups, and were consistently stronger for visceral adiposity than other measures of obesity. Central obesity alone, defined by raised waist circumference, accounted for more than half of diabetes cases in our population sample. The contribution of central obesity to diabetes prevalence was similar with cross-sectional cohorts comprising individuals from large Chinese and Indian cities^28^, but higher than amongst Chinese population samples inclusive of rural dwellers^29^.

Pathophysiological studies have produced evidence for the connection between visceral adiposity and cardiometabolic dysfunction. Imbalanced adipocytokine secretion from VAT affects the glucose-insulin homeostasis in neighboring organs^30,31^, visceral adipocyte turnover influences circulating levels of fatty-acids^30,31^, and excess VAT exerts physical compression on kidneys, increasing natriuresis and activating the sympathetic nervous system resulting in hypertension to maintain sodium balance^32^. Our findings are also consistent with the results of Mendelian Randomization studies, which support a causal effect of visceral fat on hypertension, cardiovascular disease, diabetes, and hypercholesterolemia^13^. Together, these observations provide strong evidence to support visceral adiposity and obesity as key intervention targets for prevention of diabetes and related metabolic disturbances in Asian populations, with the potential for substantial health gains.

We found that while visceral adiposity fully explained ethnic difference in triglycerides and blood pressure, it did not explain the wide differences in diabetes, insulin resistance, HDL-C, and metabolic syndrome between Indian and Chinese populations. These findings are consistent with results for studies of Indian and other South Asians in diverse settings, which consistently show an increase in diabetes and insulin resistance amongst Indian Asians, that cannot be accounted for by waist circumference^7,33^. As HOMA-IR modelling assumes similar hepatic and peripheral insulin sensitivity^17^, it is possible that unaccounted differences in hepatic insulin sensitivity might contribute to greater insulin resistance in Indian Asians^34^. Hepatic fat levels are reported to be higher in South Asians than Europeans^35^, but the contribution of hepatic insulin sensitivity towards diabetes, insulin resistance, and related metabolic disturbances in Asian populations remains to be determined. Our findings also suggest other unaccounted risk factors, which might include developmental insults^26,36^.

Our study has some limitations. Because our dataset is cross-sectional, we cannot exclude the possibility that the relationships between adiposity and cardiometabolic risk may be impacted by confounding through reverse causation. To mitigate against this risk, we carried out analyses of the relationships between adiposity and cardiometabolic risk not only in the whole study population, but also amongst people without known cardiometabolic disease. As in any population health cohort, a selective bias towards recruitment of healthier individuals is possible; however, our participants have similar characteristics to other nationally representative studies in Singapore^27,33,37^, arguing against an important healthy cohort effect. We also note that the DEXA algorithm to quantify adiposity has been validated amongst Chinese and Indian women, but has not been systematically evaluated in all the population groups we studied. Although we did not account for genetic predisposition, we note that large-scale GWAS found little evidence to support the view that genetic heterogeneity contributes to the ethnic differences in diabetes, glycemic^8^, lipid^9^, or blood pressure traits^10^. We also did not adjust our analyses for lifestyle factors such as dietary intakes and physical activity levels, modifiable exposures that impact cardiometabolic risk. Malay and Indian subgroups were reported to consume higher total calories than Chinese in Singapore^38^, but Chinese subgroups also had highest sitting time^39^ Whether unfavorable lifestyle factors contribute to ethnic differences in cardiometabolic health independently from adiposity remains unclear^40^.

Our study has several strengths. We included people of Chinese, Malay, and Indian background, making our findings relevant to a wide spectrum of global Asian populations. Participants were living in a shared environment and assessed using standardized methodologies. This design facilitates the comparison of health between populations, and reduces confounding driven by environmental or technical factors. Our analyses of continuous traits were carried out amongst people not on medication for diabetes, hypertension, and hypercholesterolemia, thus avoiding confounding by treatment effects. Finally, as conventional anthropometric measures may underestimate the impact of visceral adiposity^41^, we used DEXA-based quantification of adiposity to provide comprehensive evaluation of visceral fat across a range of indices. This approach enables quantification of fat mass across body depots and separation between visceral adiposity and overall body fat.

## Conclusion

Excess visceral adiposity has widespread impact on metabolic health in Asian populations; it increases the risk of diabetes, and is a major contributor to insulin resistance and related metabolic disturbances in 3 major Asian ethnic groups. Our results support the role of visceral adiposity as an independent risk factor for metabolic diseases, and provide strong justification for strategies to reduce excess adiposity in all Asian individuals. However, adiposity only partially explains the differences in cardiometabolic risk between Asian ethnic groups, and in particular, does not explain the high prevalence of diabetes and insulin resistance in South Asian participants. Our results thus provide the motivation to widen the search for the mechanisms responsible for divergent cardiometabolic outcomes amongst Asian populations.

## Data Availability

All data produced in the present study are available upon reasonable request to the authors

## Acknowledgement

We thank all study participants and our radiographers for help in the scanning and analysis of DEXA. We also thank former and current members of the HELIOS study steering committee, operational study team, biobanking team, and administrative team. HELIOS was supported by intramural funding from Nanyang Technological University, Lee Kong Chian School of Medicine, and the National Healthcare Group. J.C. received support from the Singapore Ministry of Health and National Medical Research Council (MOH-NMRC) STaR funding scheme [NMRC/StaR/0028/2017], LCG funding [MOH-000271], and Phase 2 National Precision Medicine Programme (Research Platform and Data Enablers) [NMRC/PRECISE/2020]. J.N. and J.L are supported by the MOH-NMRC Clinician-Scientist Awards (MOH-000654 & NMRC/CSAINV17nov005). T.M. received postdoctoral fellowship from the Lee Kong Chian School of Medicine. T.M. and J.C. had full access to all the data in this study and takes responsibility for the integrity of the data and the accuracy of the data analysis.

## Data Sharing Statement

Any data access request proposals should be directed to for the consideration of the HELIOS Study’s principal investigators.

## References

1 NCD Risk Factor Collaboration. Worldwide trends in diabetes since 1980: a pooled analysis of 751 population-based studies with 4.4 million participants. Lancet 2016; 387: 1513–30.

2 NCD Risk Factor Collaboration. Repositioning of the global epicentre of non-optimal cholesterol. Nature 2020; 582: 73–7.

3 NCD Risk Factor Collaboration. Worldwide trends in hypertension prevalence and progress in treatment and control from 1990 to 2019: a pooled analysis of 1201 population-representative studies with 104 million participants. The Lancet 2021; 398: 957–80.

4 International Diabetes Federation. IDF Diabetes Atlas (10th Edition). 2021.

5 International Diabetes Federation. IDF Diabetes Atlas (9th Edition). 2019.

6 Global Burden of Disease Collaborative Network. Global Burden of Disease Study 2019 (GBD 2019) Results. 2019. https://vizhub.healthdata.org/gbd-results/ (accessed May 17, 2023).

7 Kanaya AM, Herrington D, Vittinghoff E, et al. Understanding the high prevalence of diabetes in u.s. southasianscomparedwithfour racial/ethnic groups: The masala and mesa studies. Diabetes Care 2014; 37: 1621–8.

8 Chen J, Spracklen CN, Marenne G, et al. The trans-ancestral genomic architecture of glycemic traits. Nat Genet 2021; 53: 840–60.

9 Kuchenbaecker K, Telkar N, Reiker T, et al. The transferability of lipid loci across African, Asian and European cohorts. Nat Commun 2019; 10. DOI:10.1038/s41467-019-12026-7.

10 Giri A, Hellwege JN, Keaton JM, et al. Trans-ethnic association study of blood pressure determinants in over 750,000 individuals. Nat Genet 2019; 51: 51–62.

11 NCD Risk Factor Collaboration. Rising rural body-mass index is the main driver of the global obesity epidemic in adults. Nature 2019; 569: 260–4.

12 Jayedi A, Soltani S, Motlagh SZT, et al. Anthropometric and adiposity indicators and risk of type 2 diabetes: systematic review and dose-response meta-analysis of cohort studies. Br Med J. 2022; 376. DOI:10.1136/bmj-2021-067516.

13 Karlsson T, Rask-Andersen M, Pan G, et al. Contribution of genetics to visceral adiposity and its relation to cardiovascular and metabolic disease. Nat Med 2019; 25: 1390–5.

14 Global Health Observatory. Prevalence of obesity among adults, BMI &GreaterEqual; 30 (age-standardized estimate) (%). World Health Organization. 2016. https://www.who.int/data/gho/data/indicators/indicator-details/GHO/prevalence-of-obesity-among-adults-bmi-=-30-(age-standardized-estimate)-(-) (accessed March 1, 2022).

15 Chan SH, Bylstra Y, Teo JX, et al. Analysis of human disease variants from ancestrally diverse Asian genomes. Nat Commun 2022; 13. 10.1038/s41467-022-34116-9 (accessed July 27, 2023).

16 Mina T, Yew YW, Ng HK, et al. Adiposity impacts cognitive function in Asian populations: an epidemiological and Mendelian Randomization study. 2023 www.healthforlife.sg.

17 Wallace TM, Levy JC, Matthews DR. Use and Abuse of HOMA Modeling. Diabetes Care 2004; 27: 1487–95.

18 Ministry of Health. Singapore Disease Burden. 2018. https://www.moh.gov.sg/resources-statistics/singapore-health-facts/disease-burden.

19 Micklesfield LK, Goedecke JH, Punyanitya M, Wilson KE, Kelly TL. Dual-energy X-ray performs as well as clinical computed tomography for the measurement of visceral fat. Obesity 2012; 20: 1109–14.

20 Neeland IJ, Grundy SM, Li X, Adams-Huet B, Vega GL. Comparison of visceral fat mass measurement by dual-X-ray absorptiometry and magnetic resonance imaging in a multiethnic cohort: the Dallas Heart Study. Nutr Diabetes 2016; 6: e221–e221.

21 Alberti KGMM, Eckel RH, Grundy SM, et al. Harmonizing the metabolic syndrome: A joint interim statement of the International Diabetes Federation Task Force on Epidemiology and Prevention; National Heart, Lung, and Blood Institute; American Heart Association; World Heart Federation; International. Circulation 2009; 120: 1640–5.

22 Frost C, Thompson SG. Correcting for Regression Dilution Bias: Comparison of Methods for a Single Predictor Variable. 2000 https://www.jstor.org/stable/2680496.

23 Scott WR, Zhang W, Loh M, et al. Investigation of genetic variation underlying central obesity amongst South Asians. PLoS One 2016; 11. DOI:10.1371/journal.pone.0155478.

24 Nazare JA, Smith JD, Borel AL, et al. Ethnic influences on the relations between abdominal subcutaneous and visceral adiposity, liver fat, and cardiometabolic risk profile: The international study of prediction of intra-abdominal adiposity and its relationship with cardiometabolic risk/intra-. American Journal of Clinical Nutrition 2012; 96: 714–26.

25 Tillin T, Hughes AD, Godsland IF, et al. Insulin resistance and truncal obesity as important determinants of the greater incidence of diabetes in Indian Asians and African Caribbeans compared with Europeans: The Southall and Brent Revisited (SABRE) cohort. Diabetes Care 2013; 36: 383–93.

26 Farmaki A-E, Garfield V, Eastwood S V, et al. Type 2 diabetes risks and determinants in second-generation migrants and mixed ethnicity people of South Asian and African Caribbean descent in the UK. Diabetologia 2022; 65: 113–27.

27 Khoo CM, Khee-Shing Leow M, Sadananthan SA, et al. Body fat partitioning does not explain the interethnic variation in insulin sensitivity among asian ethnicity: the singapore adults metabolism study. Diabetes 2014; 63: 1093–102.

28 He L, Tuomilehto J, Qiao Q, et al. Impact of classical risk factors of type 2 diabetes among Asian Indian, Chinese and Japanese populations. Diabetes Metab 2015; 41: 401–9.

29 Xue H, Wang C, Li Y, et al. Incidence of type 2 diabetes and number of events attributable to abdominal obesity in China: A cohort study. J Diabetes 2016; 8: 190–8.

30 González-Muniesa P, Mártinez-González MA, Hu FB, et al. Obesity. Nat Rev Dis Primers 2017; 3. DOI:10.1038/nrdp.2017.34.

31 Karpe F, Pinnick KE. Biology of upper-body and lower-body adipose tissue - Link to whole-body phenotypes. Nat Rev Endocrinol 2015; 11: 90–100.

32 Hall JE, Do Carmo JM, Da Silva AA, Wang Z, Hall ME. Obesity-Induced Hypertension: Interaction of Neurohumoral and Renal Mechanisms. Circ Res 2015; 116: 991–1006.

33 Gao H, Salim A, Lee J, Tai ES, Van Dam RM. Can body fat distribution, adiponectin levels and inflammation explain differences in insulin resistance between ethnic Chinese, Malays and Asian Indians. Int J Obes 2012; 36: 1086–93.

34 Narayan KMV, Kanaya AM. Why are South Asians prone to type 2 diabetes? A hypothesis based on underexplored pathways. Diabetologia 2020; 63: 1103–9.

35 Iliodromiti S, McLaren J, Ghouri N, et al. Liver, visceral and subcutaneous fat in men and women of South Asian and white European descent: a systematic review and meta-analysis of new and published data. Diabetologia 2023; 66: 44–56.

36 Wells JCK. Ethnic variability in adiposity, thrifty phenotypes and cardiometabolic risk: addressing the full range of ethnicity, including those of mixed ethnicity. Obes Rev 2012; 13 Suppl 2: 14–29.

37 Khoo CM, Sairazi S, Taslim S, et al. Ethnicity modifies the relationships of insulin resistance, inflammation, and adiponectin with obesity in a multiethnic Asian population. Diabetes Care 2011; 34: 1120–6.

38 Whitton C, Rebello SA, Lee J, Tai ES, van Dam RM. A healthy asian a posteriori dietary pattern correlates with a priori dietary patterns and is associated with cardiovascular disease risk factors in a multiethnic asian population. Journal of Nutrition 2018; 148: 616–23.

39 Uijtdewilligen L, Yin JDC, van der Ploeg HP, Müller-Riemenschneider F. Correlates of occupational, leisure and total sitting time in working adults: Results from the Singapore multi-ethnic cohort. International Journal of Behavioral Nutrition and Physical Activity 2017; 14: 1–15.

40 Whitton C, Rebello SA, Lee J, Tai ES, van Dam RM. A healthy asian a posteriori dietary pattern correlates with a priori dietary patterns and is associated with cardiovascular disease risk factors in a multiethnic asian population. Journal of Nutrition 2018; 148: 616–23.

41 Vasan SK, Osmond C, Canoy D, et al. Comparison of regional fat measurements by dual-energy X-ray absorptiometry and conventional anthropometry and their association with markers of diabetes and cardiovascular disease risk. Int J Obes 2018; 42: 850–7.

