## supplementary materials for "Adiposity and metabolic health in Asian populations: An epidemiological study using Dual X-Ray Absorptiometry"

**Supplementary materials – online only**

Supplementary method

Supplementary method 1 R packages used for specific analysis or visualisation in the study

The R packages *emmeans*^1^ version 1.8.3 and *sJplot*^2^ version 2.8.12 were used to derive estimated marginal means. *ComplexHeatmap*^3^ version 2.15.1 was applied to visualize and combine multiple heatmaps. *Lavaan*^4^ version 0.6-12 was used to perform factor analysis. *AF*^5^ version 0.1.5 was applied to calculate adjusted population attribution risks.

Supplementary method 2 Repeat sub-cohort in HELIOS Study

As part of reproducibility exercise of the HELIOS Study, we invited a minimum of 240 participants (~2.4% of the total 10,004 participants) within 20 months from their first study visit to repeat the entire study protocol. The sample number was based a sample calculation to achieve a range of estimated correction factor, including from $\hat{\lambda}$=1.1 to $\hat{\lambda}$=1.5^6^. 244 completed the repeat visit, and there were 235 dataset (96.3%) with complete DEXA assessments to derive correction factor $\hat{\lambda}$, and 230 datasets (97.9%) are part of the main analysis. The mean (SD) time gap between first visit and repeat visit was 10.8 (3.4) months. In comparison, the UK Biobank invited about 4% of the total cohort, with a recall time gap ranging from 34 to 56 months^7^.

Supplementary method 3 Deriving correction factor $\hat{\boldsymbol{\lambda}}$ to correct for regression dilution bias and regression calibration

To derive correction factor $\hat{\lambda}$, we calculated Intraclass Coefficient (ICC) for BMI and vFMI as the main exposure variables for adiposity. In this study we included 3 main covariates: sex, age, and ethnicity. This is because there were no changes in sex and ethnicity assignment over time, and since age represents true aging between the baseline visit and repeat visit. As previously described, $\hat{\lambda}$= 1/ICC^8^.

If $\hat{\beta}$ represents standardized regression coefficient derived from the main analysis, and $\hat{\beta}^{*}$ represents corrected standardized regression coefficient after accounting for regression dilution bias, $\hat{\beta}^{*}=\hat{\beta}\hat{\lambda}$. Subsequently $\hat{\beta}^{*}$ and log odds ratio (OR) for the univariate regressions linking adiposity and metabolic disease and phenotype parameters could be calculated. To derive 95% confidence interval (CI) as defined by Frost and Thompson^8^:

$\hat{\beta}^{*}=\frac{f_{1}\pm\sqrt{\left( {f_{1}}^{2}-f_{0}f_{2} \right)}}{f_{2}}$	[1]

where

$f_{0}=\hat{\beta}^{2}-{1.96}^{2}var\left( \hat{\beta} \right)$	[2]

$f_{1}=\frac{\hat{\beta}}{\hat{\lambda}}-{1.96}^{2}cov\left( \hat{\beta},\frac{1}{\lambda} \right)$	[3]

$f_{2}=\frac{1}{\hat{\lambda}^{2}}-{1.96}^{2}var\left( \frac{1}{\lambda} \right)$	[4]

Since the repeat sample is a small subset of the main sample, $cov\left( \hat{\beta},\frac{1}{\lambda} \right)$ in equation [3] is close to 0 and the parameters can be treated as independent^8^. In equation [4], $var\left( \frac{1}{\lambda} \right)\approx\frac{var\left( \hat{\lambda} \right)}{\hat{\lambda}^{4}}$, and $var\left( \hat{\lambda} \right)\approx\frac{\left( \hat{\lambda}^{2}-1 \right)^{2}}{n}$.

To obtain ethnic-specific corrected delta Estimated Marginal Mean (ΔEMM) and log OR, ethnicity is assumed to be a confounder. In the presence of confounders, the ICC methods above is not applicable. Instead, we first regressed the repeat measure of the exposure on the initial measure and include sex, age, ethnicity to obtain a first-stage regression model. We then used the coefficients from the first-stage regression model to predict corrected values of the exposure for the entire dataset. We finally performed the linear or logistic regressions to obtain a second-stage regression model^7^. The second- stage regression model is thus corrected from regression dilution bias^7^, and provides $\hat{\beta}^{*}$ for the calculation of ethnic-specific corrected ΔEMM and log OR.

References:

1	Lenth R V, Bolker B, Buerkner P, *et al.* emmeans: Estimated Marginal Means, aka Least-Squares Means. https://CRAN.R-project.org/package=emmeans.

2	Ludecke D, Bartel A, Schwemmer C, Powell C, Djalovski A, Titz J. sjPlot: Data Visualization for Statistics in Social Science. https://CRAN.R-project.org/package=sjPlot.

3	Gu Z, Eils R, Schlesner M. Complex heatmaps reveal patterns and correlations in multidimensional genomic data. *Bioinformatics* 2016; **32**: 2847–9.

4	Rosseel Y. lavaan: An R Package for Structural Equation Modeling. *J Stat Softw* 2012; **48**: 1–36.

5	Dahlqwist E, Magnusson PKE, Pawitan Y, Sjölander A. On the relationship between the heritability and the attributable fraction. *Hum Genet* 2019; **138**: 425–35.

6	Morgan KE, Cook S, Leon DA, Frost C. Reflection on modern methods: Calculating a sample size for a repeatability sub-study to correct for measurement error in a single continuous exposure. *Int J Epidemiol* 2019; **48**: 1721–6.

7	Rutter CE, Millard LA, Borges MC, Lawlor DA. Exploring regression dilution bias using repeat measurements of 2858 variables in up to 49 000 UK Biobank participants. *medRxiv* 2022. DOI:10.1101/2022.07.13.22277605.

8	Frost C, Thompson SG. Correcting for Regression Dilution Bias: Comparison of Methods for a Single Predictor Variable. 2000 https://www.jstor.org/stable/2680496.

| Supplementary Table 1. A comparison of the HELIOS Study with the Singapore National Statistics | | | |
| --- | --- | --- | --- |
|  | | **Singapore** ^1^ | **HELIOS dataset for this manuscript**, n=9067^2^ |
| **Sex** | Male: Female | 1:1.1 | 1: 1.5 |
| **Ethnicity** | Chinese: Malay: Indian | 76:15:9 | 69:13:18 |
| **Age** (years) | | 40.8* | 52.82 ± 11.76 |
| **Lifestyle** | Current smoking, % | 12 | 8.4 |
| **Education** | Total Year of Education (years) | 11.1 | 13.80 ± 3.44 |
| **Overall health** | Diabetes (Glucose ≥ 7.0 mmol/L), % | 8.6 | 8.3 |
|  | Hypertension (SBP/DBP ≥ 140/90 mmHg), % | 21.5 | 18.0 |
|  | Hypercholesterolemia (LDL≥ 4.100 mmol/L), % | 33.6 | 29.0 |
|  | Obesity (BMI ≥ 30.0 Kg/m^2^), % | 8.7 | 12.2 |
| ^1^ Data were taken from the Population Trend 2018, Singapore Department of Statistics, and the Singapore Disease Burden 2017 for adults aged 18 to 69 years, Ministry of Health (<https://www.moh.gov.sg/resources-statistics/singapore-health-facts/disease-burden>).  **^2^** This refers to the dataset before the removal of persons with medication for diabetes, hypertension, and hypercholesterolemia.  *The national data were presented in median only for all age groups include children. | | | |

| Supplementary Table 2 Adjusted Population Attributable Risk Proportion of adiposity on cardiometabolic diseases.  Values are expressed as percentage (%) with 95% confidence interval (CI). Covariates include sex, age, and ethnicity (in the overall pool only). Overweight and Obesity are defined as BMI ≥23.0 Kg/m^2^ and >27.5 Kg/m^2^, respectively. Central obesity is defined as ≥90 cm and ≥80 cm for male and female, respectively, in line with the metabolic syndrome definition by International Diabetes Federation and American Heart Association 2009. | | | |
| --- | --- | --- | --- |
| **Adiposity exposure** | **Diabetes** | **Hypertension** | **Hypercholesterolemia** |
| **Overall, n=9067** |  |  |  |
| Overweight | 30.8 (21.5: 40.2) | 45.6 (40.8: 50.4) | 17.6 (13.5: 21.6) |
| Obesity | 16.0 (11.2: 20.8) | 19.9 (17.2: 22.6) | 4.9 (3.1: 6.8) |
| Central obesity | 40.8 (34.4: 47.3) | 34.5 (30.7: 38.3) | 13.4 (10.5: 16.3) |
| **Chinese, n=6224** |  |  |  |
| Overweight | 37.5 (26.8: 48.2) | 42.9 (37.7: 48.1) | 17 (12.9: 21) |
| Obesity | 17.6 (12.0: 23.2) | 15.1 (12.4: 17.8) | 3.9 (2.3: 5.6) |
| Central obesity | 40.5 (32.5: 48.5) | 30.8 (26.8: 34.9) | 11.4 (8.5: 14.2) |
| **Malay, n=1169** |  |  |  |
| Overweight | 70.5 (48.1: 92.9) | 66 (46.6: 85.4) | 11.6 (-7.8: 31.1) |
| Obesity | 30.8 (17.2: 44.4) | 39.0 (28.8: 49.4) | 13.3 (0.6: 20.8) |
| Central obesity | 46.8 (30: 63.6) | 54.1 (41.5: 66.8) | 18.3 (7.8: 28.8) |
| **Indian, n=1674** |  |  |  |
| Overweight | -8.1 (-28.7: 12.5) | 37 (22.3: 51.7) | 14.4 (1.1: 27.7) |
| Obesity | 0.3 (-0.6: 10.7) | 18.9 (12.1: 25.7) | 1.4 (-0.4: 0.6) |
| Central obesity | 30.7 (17.6: 43.8) | 26.8 (15.7: 37.9) | 13.8 (4.6: 23) |

| Supplementary Table 3 Visceral adiposity, HOMA-IR, and diabetes status across Asian BMI categories. vFMI and waist circumference were expressed as mean ± SD. HOMA-IR is presented as geometric mean ± SD. The diabetes status was expressed in percentage (%). vFMI= visceral Fat Mass Index; HOMA-IR=Homeostatic Model Assessment for Insulin Resistance. | | | | |
| --- | --- | --- | --- | --- |
| **Cardiometabolic factors** | **Ethnicity** | **BMI (Kg/m2)** | | |
|  |  | **<23** | **23-27.5** | **>27.5** |
| **vFMI**, Kg/m^2^ | Chinese | 0.15 ± 0.06 | 0.23 ± 0.06 | 0.33 ± 0.09 |
|  | Malay | 0.14 ± 0.05 | 0.23 ± 0.07 | 0.34 ± 0.09 |
|  | Indian | 0.17 ± 0.06 | 0.25 ± 0.06 | 0.34 ± 0.09 |
| **waist circumference**, cm | Chinese | 72.94 ± 6.55 | 83.1 ± 6.64 | 94.54 ± 9.03 |
|  | Malay | 72.89 ± 6.87 | 82.18 ± 6.96 | 94.22 ± 9.45 |
|  | Indian | 75.79 ± 7.11 | 86.07 ± 7.38 | 97.3 ± 9.82 |
| **HOMA-IR** | Chinese | 1.16 ± 0.76 | 1.8 ± 1.26 | 3.16 ± 2.75 |
|  | Malay | 1.17 ± 0.91 | 1.72 ± 1.30 | 2.9 ± 2.26 |
|  | Indian | 1.52 ± 1.11 | 2.3 ± 1.82 | 3.69 ± 3.05 |
| **Diabetes** | Chinese | 3 | 5.6 | 10 |
|  | Malay | 3.7 | 11.7 | 17.8 |
|  | Indian | 18.7 | 17.5 | 17.2 |

| Supplementary Table 4. Odds ratio of disease outcomes before and after adjusted for adiposity indices. vFMI= visceral Fat Mass Index | | | | | |
| --- | --- | --- | --- | --- | --- |
|  | **Ethnicity** | **Adjusted for sex and age** | **Adjusted for sex, age, BMI** | **Adjusted for sex, age, waist circumference** | **Adjusted for sex, age, vFMI** |
| Diabetes | Malay | 3.7 [3; 4.6] | 2 [1.6; 2.5] | 2.3 [1.8; 2.8] | 2.2 [1.8; 2.8] |
|  | Indian | 4.3 [3.6; 5.1] | 3.2 [2.7; 3.9] | 2.8 [2.3; 3.3] | 3 [2.5; 3.6] |
| Hypercholesterolemia | Malay | 1.7 [1.5; 2] | 1.2 [1.1; 1.4] | 1.4 [1.2; 1.6] | 1.3 [1.1; 1.5] |
|  | Indian | 1.3 [1.1; 1.4] | 1 [0.9; 1.2] | 1 [0.9; 1.1] | 1 [0.9; 1.1] |
| Hypertension | Malay | 1.5 [1.3; 1.8] | 0.7 [0.6; 0.8] | 0.9 [0.7; 1.1] | 0.9 [0.7; 1.1] |
|  | Indian | 1.5 [1.3; 1.7] | 0.9 [0.8; 1] | 0.9 [0.7; 1] | 0.9 [0.8; 1.1] |

| Supplementary Table 5. EMM±SEM of metabolic parameters before and after adjusted for adiposity indices in SI unit. Regressions were performed in a subset of participants with no medication for diabetes, hypertension, and hypercholesterolemia, n=6,807. Geometric mean ± SEM were presented for glucose, insulin, HOMA-IR, HOMA-%B, triglycerides, and HDL-C. | | | | |
| --- | --- | --- | --- | --- |
| **Metabolic parameters** | **Ethnicity** | **Adjusted for sex and age** | **Adjusted for sex, age, BMI** | **Adjusted for sex, age, vFMI** |
| Glucose (mmol/L) | Chinese | 4.8 ± 0.02 | 4.84 ± 0.02 | 4.84 ± 0.02 |
|  | Malay | 4.94 ± 0.04 | 4.82 ± 0.04 | 4.84 ± 0.04 |
|  | Indian | 5.04 ± 0.04 | 4.95 ± 0.04 | 4.94 ± 0.04 |
| Insulin (μIU/mL) | Chinese | 7.25 ± 0.03 | 7.87 ± 0.02 | 7.83 ± 0.02 |
|  | Malay | 9.47 ± 0.06 | 7.25 ± 0.05 | 7.83 ± 0.05 |
|  | Indian | 11.38 ± 0.05 | 9.4 ± 0.05 | 9.44 ± 0.05 |
| HOMA-IR | Chinese | 1.55 ± 0.03 | 1.69 ± 0.02 | 1.68 ± 0.02 |
|  | Malay | 2.08 ± 0.07 | 1.55 ± 0.06 | 1.68 ± 0.06 |
|  | Indian | 2.55 ± 0.06 | 2.07 ± 0.05 | 2.07 ± 0.05 |
| HOMA-%B | Chinese | 117.43 ± 0.02 | 124.24 ± 0.02 | 123.52 ± 0.02 |
|  | Malay | 147.15 ± 0.06 | 122.62 ± 0.06 | 129.78 ± 0.06 |
|  | Indian | 163.3 ± 0.06 | 143.42 ± 0.05 | 144.46 ± 0.05 |
| Triglycerides (mmol/L) | Chinese | 1.04 ± 0.02 | 1.08 ± 0.02 | 1.09 ± 0.02 |
|  | Malay | 1.22 ± 0.05 | 1.07 ± 0.05 | 1.09 ± 0.05 |
|  | Indian | 1.2 ± 0.05 | 1.1 ± 0.04 | 1.08 ± 0.04 |
| HDL-C (mmol/L) | Chinese | 1.55 ± 0.02 | 1.52 ± 0.02 | 1.52 ± 0.02 |
|  | Malay | 1.36 ± 0.04 | 1.46 ± 0.04 | 1.44 ± 0.04 |
|  | Indian | 1.27 ± 0.04 | 1.34 ± 0.04 | 1.34 ± 0.04 |
| Systolic BP (mmHg) | Chinese | 118.71 ± 0.23 | 119.8 ± 0.23 | 119.67 ± 0.23 |
|  | Malay | 121.59 ± 0.54 | 118.06 ± 0.54 | 119.23 ± 0.53 |
|  | Indian | 119.69 ± 0.47 | 117.17 ± 0.46 | 117.38 ± 0.46 |
| Diastolic BP (mmHg) | Chinese | 70.49 ± 0.15 | 71.11 ± 0.15 | 71.06 ± 0.14 |
|  | Malay | 73.81 ± 0.34 | 71.83 ± 0.34 | 72.41 ± 0.33 |
|  | Indian | 73.44 ± 0.29 | 72.02 ± 0.29 | 72.06 ± 0.29 |

| Supplementary Table 6. Δ Estimated Marginal Means ±SEM of glycemic indices in Indian participants (compared to Chinese as reference) before and after adjusting for various adiposity indices. All univariate regressions were adjusted for sex and age, and ethnicity. Glucose, insulin, HOMA-IR and HOMA-%B were all ln-transformed and z-scored. | | | | |
| --- | --- | --- | --- | --- |
| **Adiposity indices as covariate** | **Glucose** | **Insulin** | **HOMA-IR** | **HOMA-%B** |
| demographic factors only | 0.34 ± 0.03 | 0.45 ± 0.03 | 0.74 ± 0.03 | 0.75 ± 0.03 |
| BMI | 0.16 ± 0.03 | 0.27 ± 0.03 | 0.29 ± 0.03 | 0.3 ± 0.03 |
| Waist circumference | 0.14 ± 0.03 | 0.27 ± 0.03 | 0.28 ± 0.03 | 0.23 ± 0.03 |
| Waist-hip ratio | 0.25 ± 0.03 | 0.58 ± 0.03 | 0.58 ± 0.03 | 0.45 ± 0.03 |
| visceral FMI | 0.14 ± 0.03 | 0.26 ± 0.03 | 0.31 ± 0.03 | 0.31 ± 0.03 |
| trunk FMI | 0.13 ± 0.03 | 0.22 ± 0.03 | 0.23 ± 0.03 | 0.19 ± 0.03 |
| Android FMI | 0.14 ± 0.03 | -0.24 ± 0.03 | 0.25 ± 0.03 | 0.2 ± 0.03 |
| Gynoid FMI | 0.22 ± 0.03 | 0.31 ± 0.03 | 0.33 ± 0.03 | 0.22 ± 0.03 |
| Total FMI | 0.14 ± 0.03 | 0.18 ± 0.03 | 0.19 ± 0.03 | 0.15 ± 0.03 |

| Supplementary Table 7. Intra-class correlations (ICC), correction factor $\hat{\boldsymbol{\lambda}}$, $\boldsymbol{var}\left( \frac{\boldsymbol{1}}{\boldsymbol{\lambda}} \right)$, and $\boldsymbol{var}\left( \hat{\boldsymbol{\lambda}} \right)$ for independent variables. Abbreviations (A-Z): BMI= body mass index; VAT=Visceral adipose tissue, vFMI= visceral Fat Mass Index. | | | | |
| --- | --- | --- | --- | --- |
| **Adiposity parameters** | **ICC (95% CI)** | $\hat{\boldsymbol{\lambda}}$ | $\boldsymbol{var}\left( \frac{\boldsymbol{1}}{\hat{\boldsymbol{\lambda}}} \right)$ | $\boldsymbol{var}\left( \hat{\boldsymbol{\lambda}} \right)$ |
| Height, cm | 1 (1; 1) | 1.00 | 4.46 x10^-8^ | 4.48 x10^-8^ |
| Weight, Kg | 0.98 (0.97; 0.98) | 1.02 | 6.84 x10^-6^ | 7.41 x10^-6^ |
| BMI, Kg/m^2^ | 0.97 (0.96; 0.98) | 1.03 | 1.36 x10^-5^ | 1.52 x10^-5^ |
| VAT, Kg | 0.94 (0.92; 0.95) | 1.06 | 5.76 x10^-5^ | 7.35 x10^-5^ |
| vFMI, Kg/m^2^ | 0.94 (0.92; 0.95) | 1.06 | 5.66 x10^-5^ | 7.21 x10^-5^ |

| Supplementary Table 8 Estimated target BMI and waist circumference for Malay and Indian participants to achieve insulin sensitivity as observed in Chinese participants | | |
| --- | --- | --- |
| **Ethnicity, sex** | **Equation for HOMA-IR ~BMI,** where mean (SD) of the overall sample = 24.4 (4.4) Kg/m^2^ | **Target BMI (Kg/m^2^)** |
| Malay, female | HOMA-IR = 2.17 -0.31*ethnic Malay + 0.00*age -0.14*female + 1.02*z-BMI | 23.5 |
| Malay, male | HOMA-IR = 2.17 -0.31*ethnic Malay + 0.00*age +0.14*male + 1.02*z-BMI | 22.3 |
| Indian, female | HOMA-IR = 2.17 +0.53*ethnic Indian + 0.00*age -0.14*female + 1.02*z-BMI | 19.9 |
| Indian, male | HOMA-IR = 2.17 +0.53*ethnic Indian + 0.00*age +0.14*male + 1.02*z-BMI | 18.7 |
|  | **Equation for HOMA-IR~waist** **circumference**, where mean (SD) of the overall sample = 81.5 (11.2) cm, and averaged age =50.19 years | **Target waist circumference (cm)** |
| Malay, female | HOMA-IR = 2.29-0.01*ethnic Malay -0.01*age + 0.45*female +1.08*z-waist | 74.2 |
| Malay, male | HOMA-IR = 2.29-0.01*ethnic Malay -0.01*age - 0.45*female +1.08*z-waist | 83.5 |
| Indian, female | HOMA-IR = 2.29+0.49*ethnic Indian -0.01*age + 0.45*female +1.08*z-waist | 69.0 |
| Indian, male | HOMA-IR = 2.29+0.49*ethnic Indian -0.01*age - 0.45*female +1.08*z-waist | 78.3 |

Supplementary Figure 1. CONSORT diagram of the dataset selection.

Orange box indices 3 analysis datasets used in this study. Dotted lines represent data subset. * Data were unavailable or incomplete due to the presence of contraindications or other operational reasons.

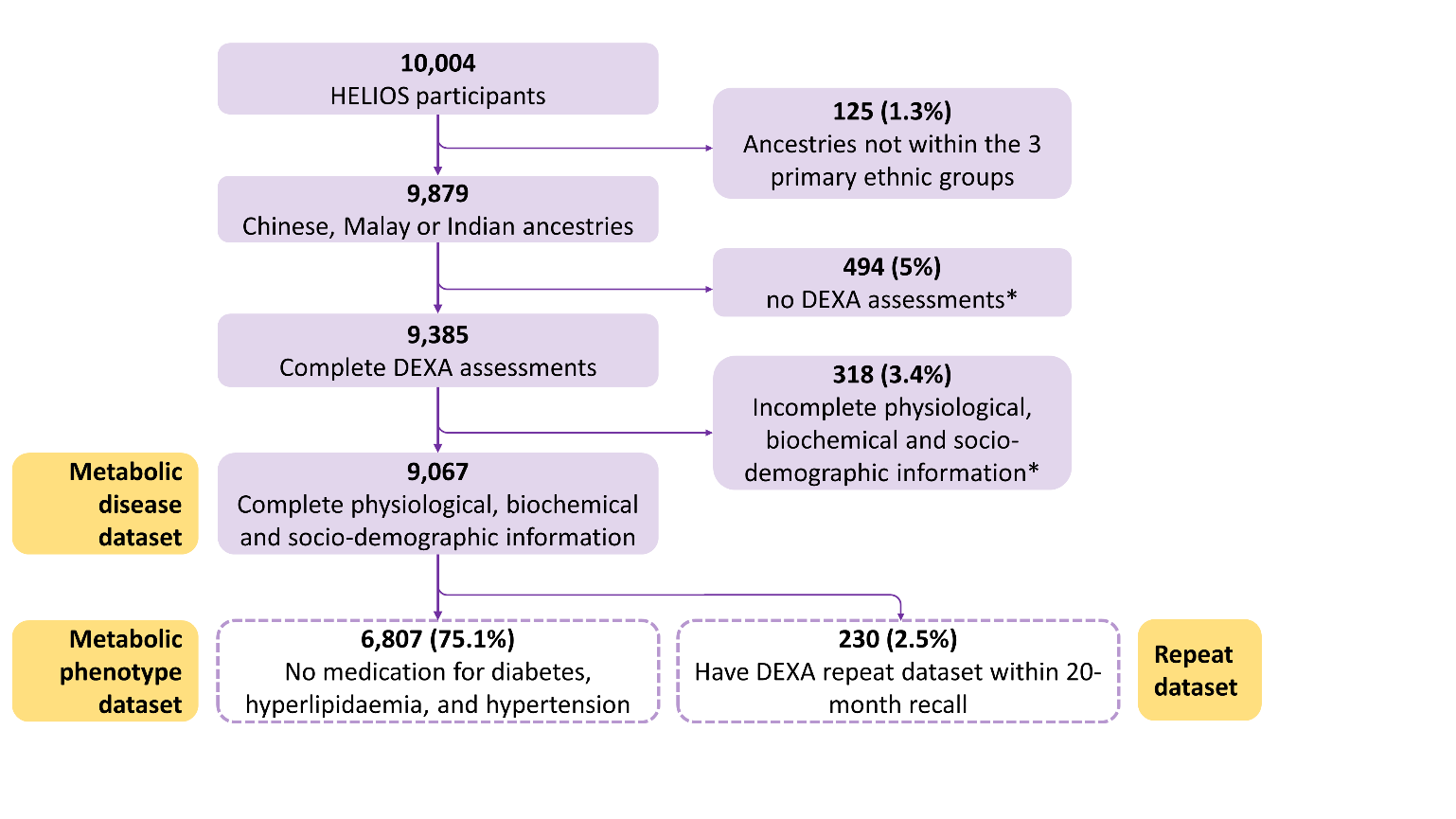

Supplementary Figure 2 Contribution of excess visceral adiposity in metabolic health.

All analyses were adjusted for sex and age, and ethnicity. **A)** Univariate logistic associations of adiposity indices with metabolic diseases (n **=** 9,067)**.** Each point is Odds Ratio (OR) with 95% Confidence Interval (CI). **B)** Univariate associations of adiposity indies with metabolic phenotypes (n=6,807)**.** Glucose, insulin, HOMA-IR, HOMA-%B, triglycerides and HDL-C were all ln-transformed and z-scored. The number in each cell is standardized beta and were all statistically significant at 0.05 levels. The associations with HDL-C were inverted. **C)** Univariate associations of adiposity indices with metabolic phenotypes in Chinese (n=4,789), Malay (n=879), and Indian Asians (n=1,139)**. Abbreviations (A-Z):** BMI= body mass index; DBP= diastolic blood pressure; HDL-C = High-density lipoproteins cholesterols; HOMA-B= Homeostatic model assessment (HOMA) of β-cell function; HOMA-IR=Homeostatic Model Assessment for Insulin Resistance; hyperchol.= hypercholesterolemia. SBP= systolic blood pressure; T2D= Type 2 diabetes; vFMI= visceral Fat Mass Index.

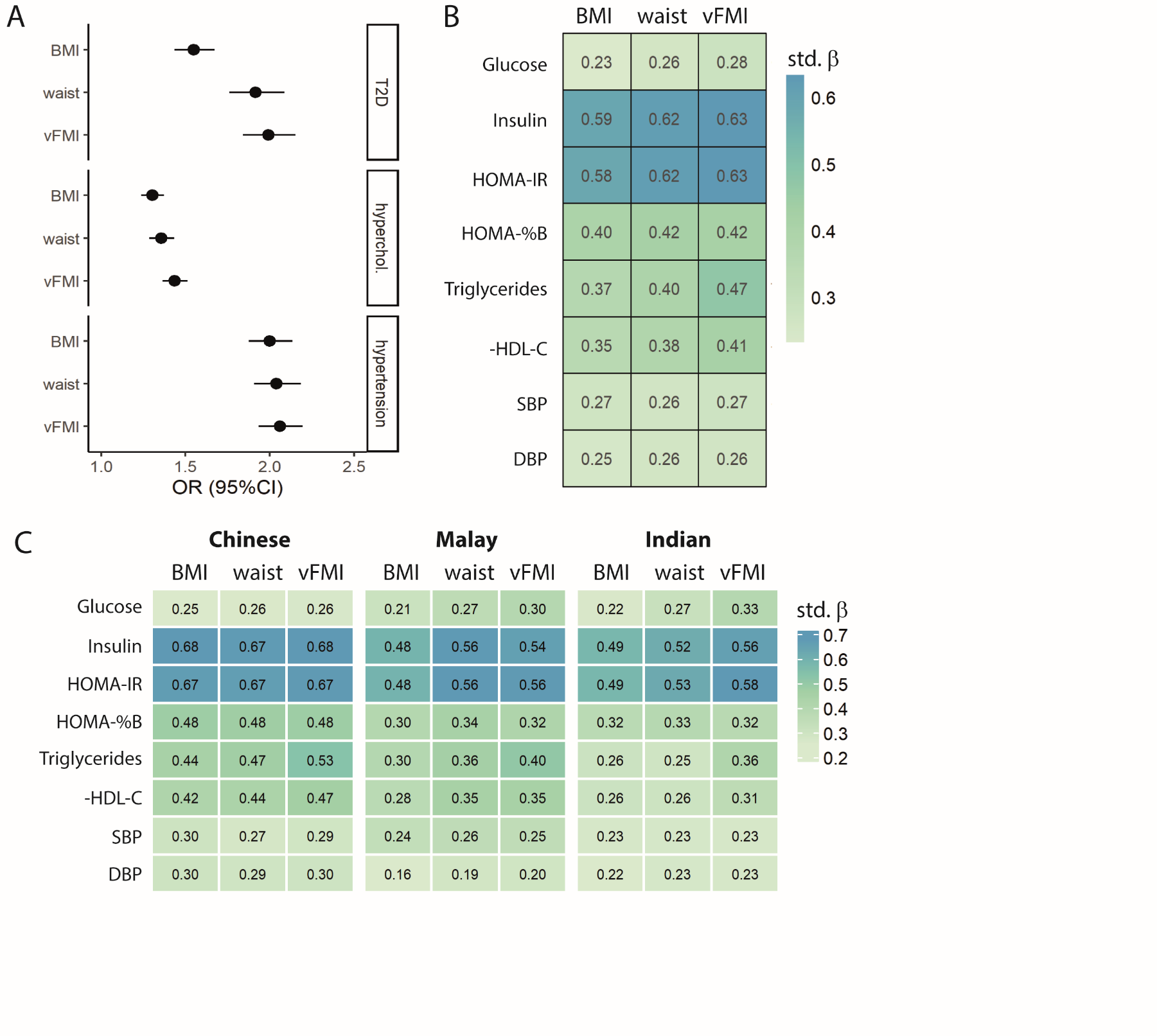

Supplementary Figure 3. Contribution of excess visceral adiposity in metabolic health across ethnicity in all eligible dataset, including participants with medication for diabetes, hypertension, and hyperlipidaemia, n= 9,067.

All analyses were adjusted for sex and age. Glucose, insulin, HOMA-IR, HOMA-%B, triglycerides and HDL-C were all ln-transformed and z-scored. Each bar represents Δ Estimated Marginal Means ±SEM of z-scores. ΔEMM for z-HDL-C was inversed to ease comparison. The 3 regression models were i) demog= adjusted for sex and age; ii) BMI=model i) + BMI; and iii) vFMI=model i) + vFMI. ** p<0.001 and * p <0.05 compared to Chinese as reference. **Abbreviations (A-Z):** BMI= body mass index; DBP= diastolic blood pressure; HDL-C = High-density lipoproteins cholesterols; HOMA-B= Homeostatic model assessment (HOMA) of β-cell function; HOMA-IR=Homeostatic Model Assessment for Insulin Resistance; SBP= systolic blood pressure; vFMI= visceral Fat Mass Index.

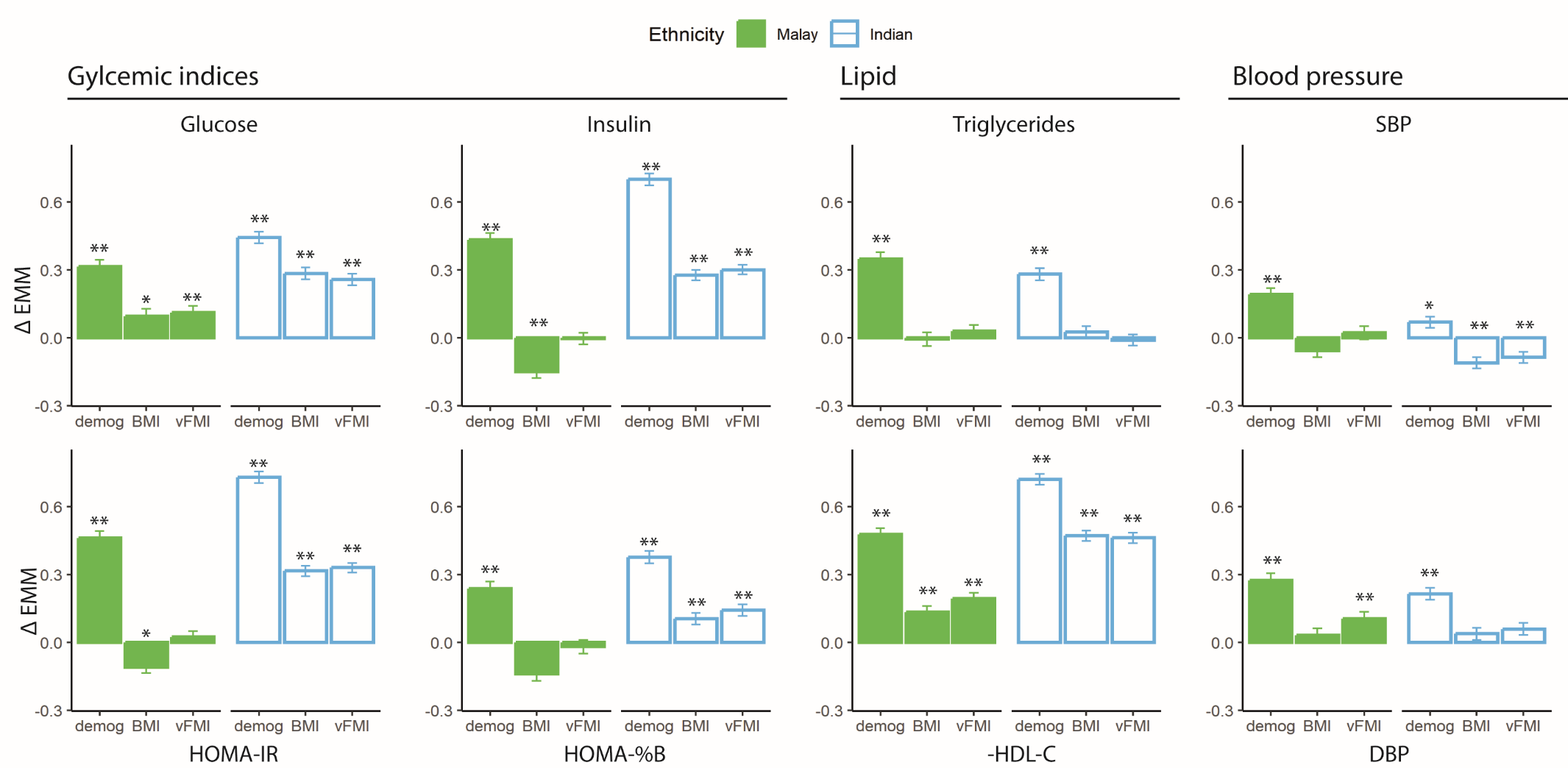

Supplementary Figure 4 Contribution of excess visceral adiposity in metabolic health across ethnicity using untransformed raw data. All analyses were adjusted for sex and age, and were performed in participants without medication for diabetes, hypertension, and hyperlipidaemia, n= 6,807. Each bar represents Δ Estimated Marginal Means ±SEM before and after adjusting for adiposity indices. ΔEMM for HDL-C was inversed to ease comparison. The 3 regression models were i) demog= adjusted for sex and age; ii) BMI=model i) + BMI; and iii) vFMI=model i) + vFMI. ** p<0.001 and * p <0.05 compared to Chinese participants as reference. Abbreviations (A-Z): BMI= body mass index; DBP= diastolic blood pressure; Gluc= Glucose; HDL-C= High-density lipoprotein cholesterol; HOMA-%B= Homeostatic model assessment (HOMA) of β-cell function; HOMA-IR=Homeostatic Model Assessment for Insulin Resistance; SBP= systolic blood pressure; vFMI= visceral Fat Mass Index. Fat mass estimated as BMI or vFMI does not explain differences between Indian and Chinese participants in glycemic indices and HDL-C levels.

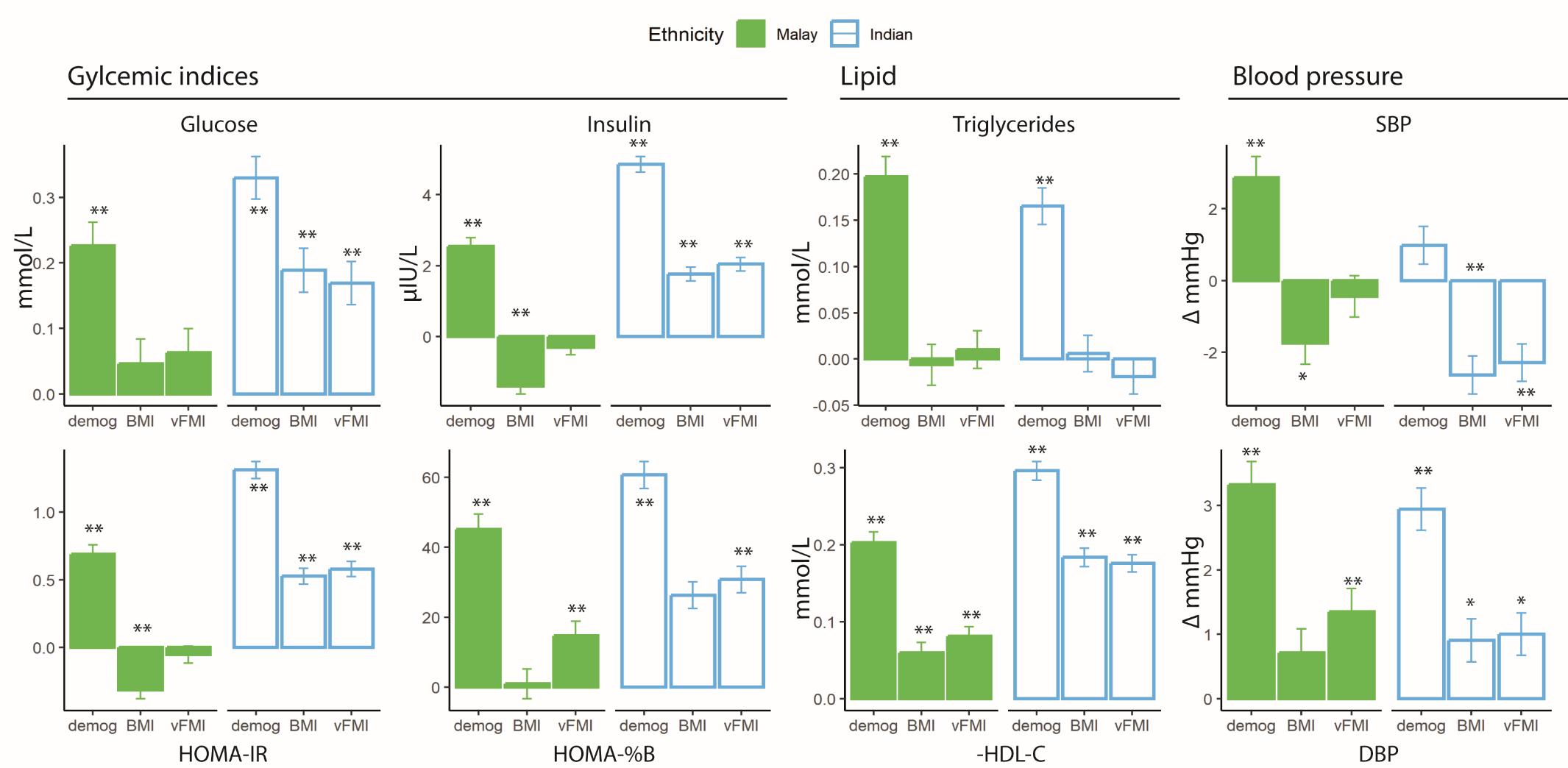

Supplementary Figure 5. Contribution of excess visceral adiposity in metabolic health across ethnicity after correction factor was introduced.

The analysis was performed amongst participants with medication for diabetes, hypertension, and hyperlipidaemia, n=6,807. All analyses were adjusted for sex and age. Glucose, insulin, HOMA-IR, HOMA-%B, triglycerides and HDL-C were all ln-transformed and z-scored. Each bar represents Δ Estimated Marginal Means ±SEM of z-scores. ΔEMM for z-HDL-C was inversed to ease comparison. The 3 regression models were i) demog= adjusted for sex and age; ii) BMI=model i) + BMI; and iii) vFMI=model i) + vFMI. ** p<0.001 and * p <0.05 compared to Chinese as reference. **Abbreviations (A-Z):** BMI= body mass index; DBP= diastolic blood pressure; HDL-C = High-density lipoproteins cholesterols; HOMA-B= Homeostatic model assessment (HOMA) of β-cell function; HOMA-IR=Homeostatic Model Assessment for Insulin Resistance; SBP= systolic blood pressure; vFMI= visceral Fat Mass Index.

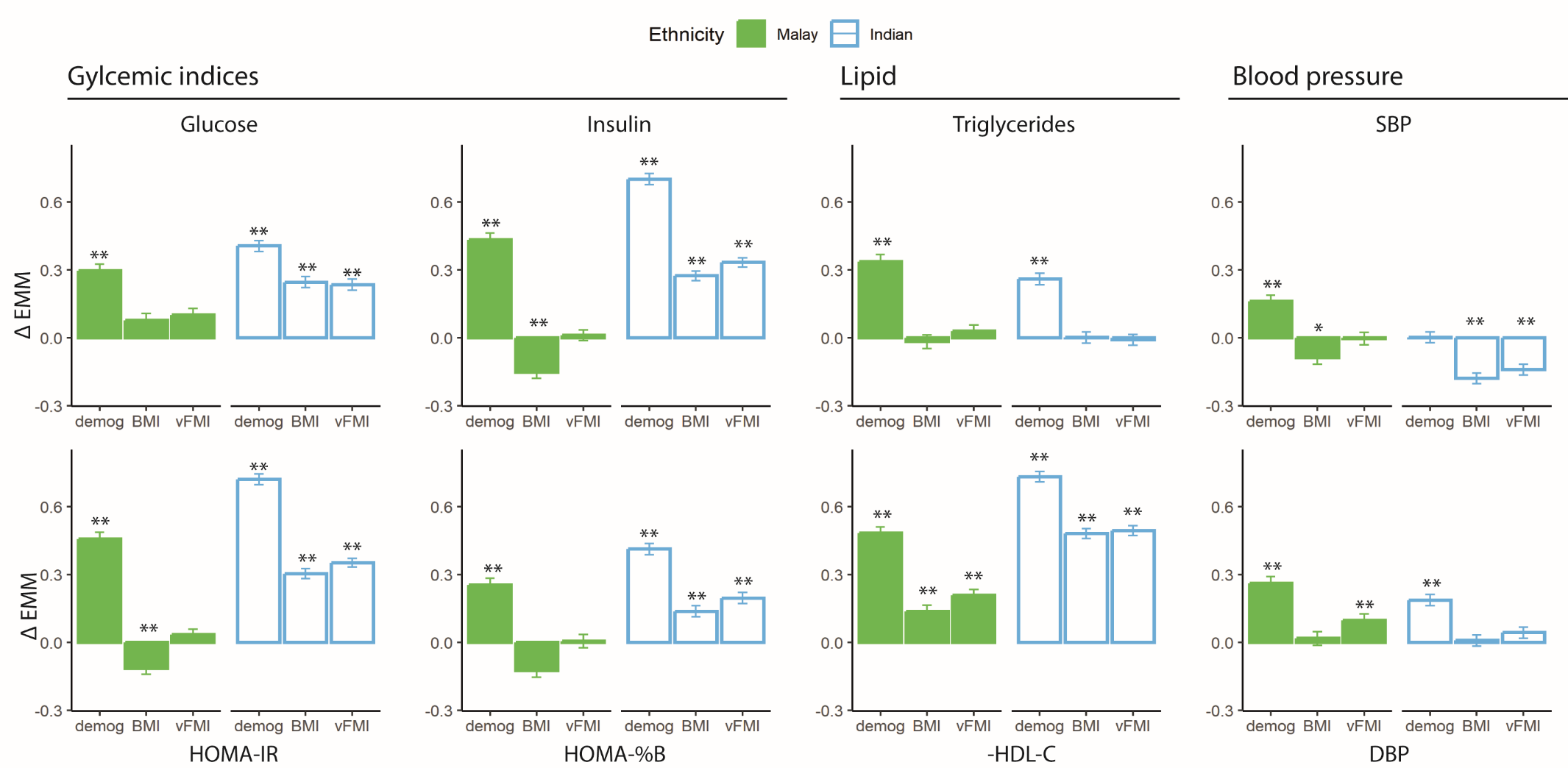

Supplementary Figure 6 Contribution of excess visceral adiposity in metabolic health across ethnicity, adjusting for socio-economic status. All analyses were performed in participants without medication for diabetes, hypertension, and hyperlipidaemia, n= 6,807. All analyses were adjusted for sex and age. Glucose, insulin, HOMA-IR, HOMA-%B, triglycerides and HDL-C were all ln-transformed and z-scored. Each bar represents Δ Estimated Marginal Means ±SEM of z-scores. ΔEMM for z-HDL-C was inversed to ease comparison. The 4 regression models were i) demog= adjusted for sex and age; ii) ses=model i) + cigarette smoking, alcohol intake, total year of education and household income; iii) vFMI=model i) + vFMI; iv) ses_vFMI=model ii) + cigarette smoking, alcohol intake, total year of education and household income.** p<0.001 and * p <0.05 compared to Chinese as reference. Abbreviations (A-Z): DBP= diastolic blood pressure; HDL-C = High-density lipoproteins cholesterols; HOMA-B= Homeostatic model assessment (HOMA) of β-cell function; HOMA-IR=Homeostatic Model Assessment for Insulin Resistance; ses= socio-economic status. SBP= systolic blood pressure; vFMI= visceral Fat Mass Index.

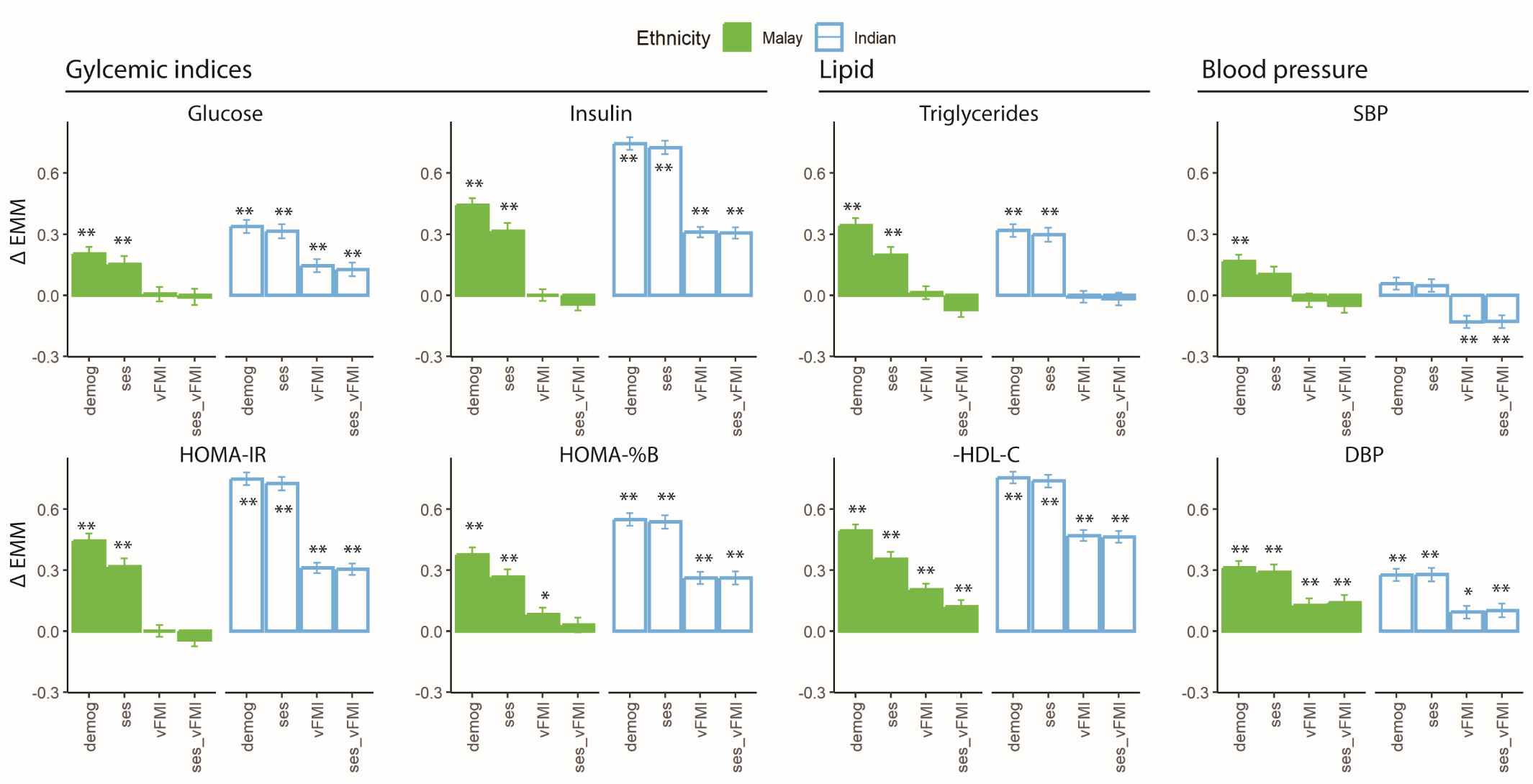

Supplementary Figure 7. Hierarchical clustering of partial correlations amongst metabolic phenotypes and adiposity indices, n=6,807.

All analyses were adjusted for sex and age, and ethnicity. Glucose, insulin, HOMA-IR, HOMA-%B, triglycerides and HDL-C were all ln-transformed and z-scored. **Abbreviations (A-Z):** BMI= body mass index; DBP= diastolic blood pressure; Gluc= Glucose; HDL-C = High- lipoproteins cholesterols; HOMA-%B= Homeostatic model assessment (HOMA) of β-cell function; HOMA-IR=Homeostatic Model Assessment for Insulin Resistance; SBP= systolic blood pressure; Trig= Triglycerides; vFMI= visceral Fat Mass Index; waist= waist circumference.

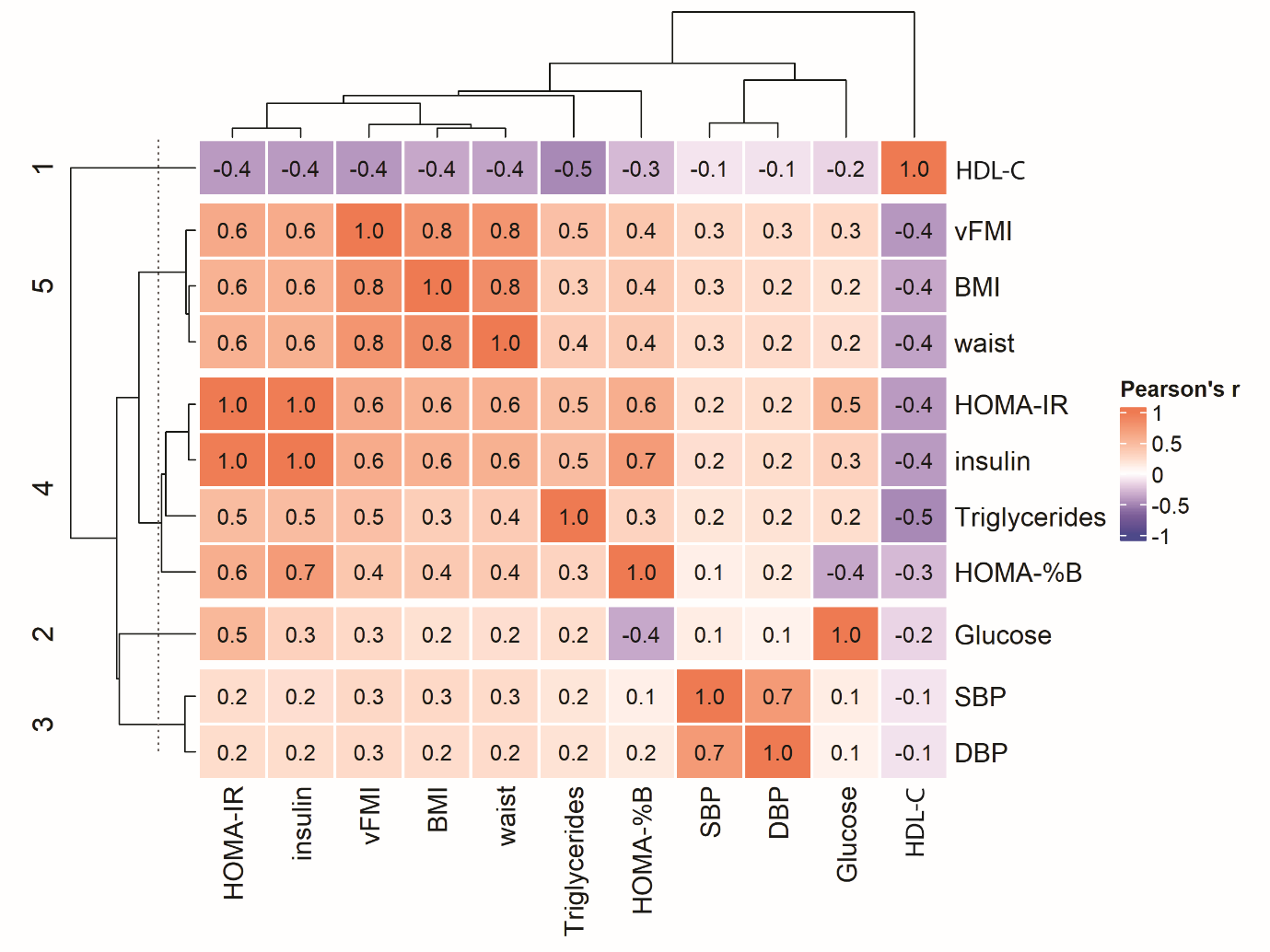

Supplementary Figure 8. Contribution of visceral adiposity in metabolic Principal Components (PCs) across ethnicity.

All analyses were adjusted for sex and age. Ethnic difference in metabolic PC before and after adjusting for adiposity measures. Each bar represents Δ Estimated Marginal Means ±SEM compared to Chinese as reference, x-axis indices covariates (sex and age), with 1 additional adiposity parameter. ** p<0.001 and * p <0.05 compared to Chinese as reference**. Abbreviations (A-Z):** BMI= body mass index; vFMI= visceral Fat Mass Index.

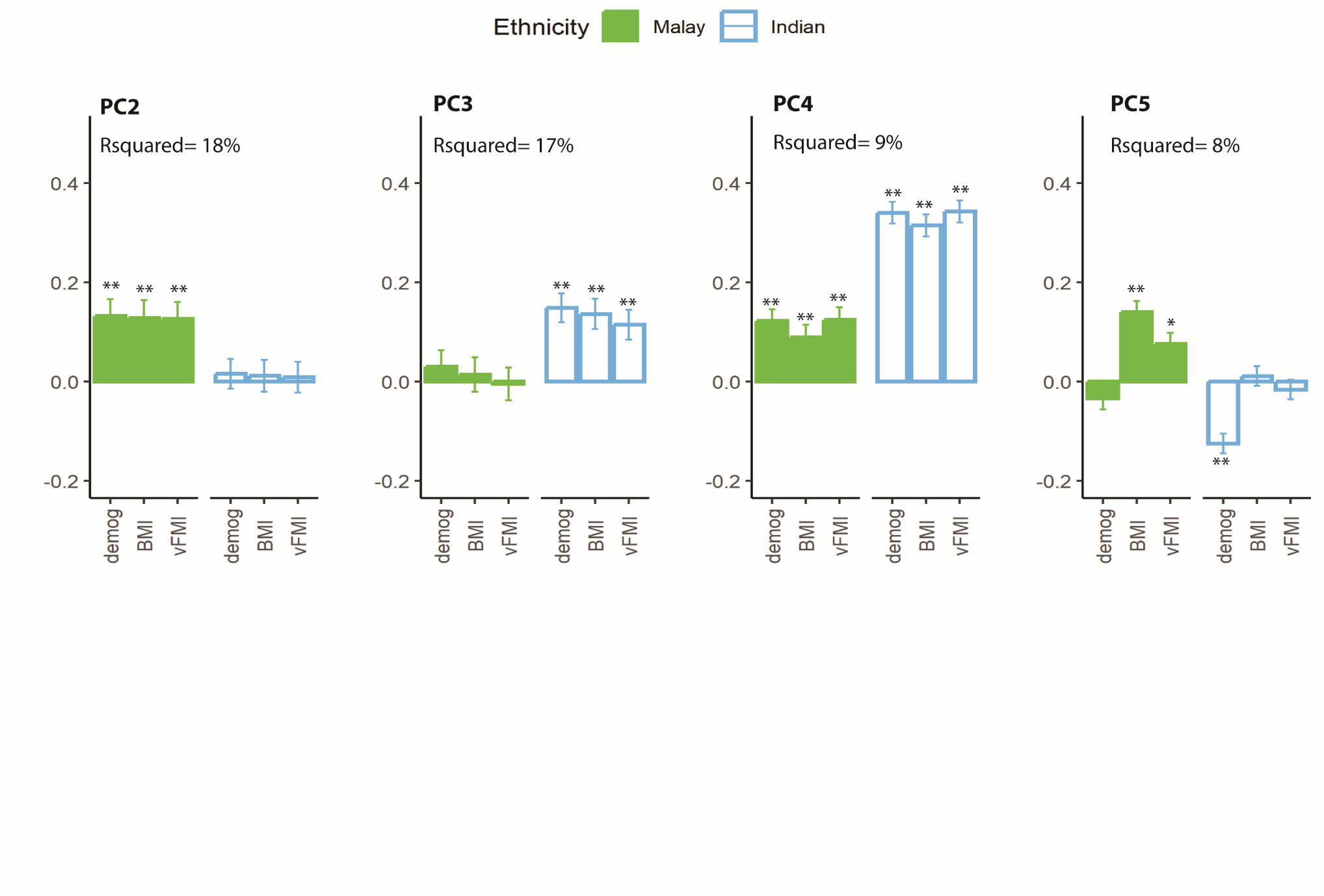
